# Understanding Sexual Violence in the Colombian Armed Conflict: Victim Characteristics, Spatial Clustering, and Temporal Contagion

**DOI:** 10.1101/2025.11.12.25340057

**Authors:** Elisavet Pappa, Alida Acosta-Ortiz, Charlotte Constable Fernandez, María Camila García Durán, Charlotte M. Jones Todd, Rob Saunders, Francesca Solmi, William Tamayo-Agudelo, Fabio Idrobo, Vaughan Bell

## Abstract

**Background:** Sexual violence is one of the most severe forms of civilian victimisation in the Colombian armed conflict. Despite its importance, population-scale analysis of conflict-related sexual violence patterns is limited, something essential for informing public health responses and prevention strategies.

**Methods:** We analysed two national databases of armed-conflict-related data from Colombia: the statutory Register of Victims and the National Centre for Historical Memory. We profiled victim demographics and used i) log-Gaussian Cox process modelling to identify geospatial clustering and ii) Hawkes process modelling to examine temporal contagion. Analyses covered the entire conflict period (1964-2024) and a recent period (2014-2024).

**Results:** Victims were predominantly female (≈90%) and disproportionately from ethnic minorities. Sexual violence showed geographic clustering, with a spatial range of up to 2.4km for the entire conflict period and reducing to 1.7km after adjustment for population density. In 2014-2024, baseline intensity decreased but spatial clustering extended over wider areas (range = 2.5km unadjusted; 1.9km adjusted). For the entire conflict, the unadjusted Hawkes model indicated near-critical temporal dependency (branching ratio = 0.99) and multi-day temporal contagion (half-life = 3.7 days). The model, adjusted for changes in background rates, revealed little evidence for temporal contagion independent of changes in background rates. For 2014-2024, the unadjusted model similarly showed near-critical branching (branching ratio = 1.00) with extended decay (half-life = 10.04 days), whereas background rate-adjusted models indicated little evidence for temporal contagion beyond background rate changes.

**Conclusions:** Sexual violence in the Colombian armed conflict may be more widely present than is apparent from raw event counts and has shifted from concentrated, high-intensity incidents to more dispersed patterns. Areas of historically high prevalence remain critical for victim support. The temporal dynamics suggest that prevention efforts may benefit from proactively addressing long-term structural and contextual determinants.

## Introduction

The Colombian armed conflict has endured for almost six decades and has disproportionally affected civilians with an estimated 18.3% of the entire population affected by at least one form of violence (Unidad para las Víctimas, 2023). Sexual violence has been identified as one of the most serious types of civilian victimisation by the Colombian transitional justice process (Lopera & Concha, 2022) and victims of conflict related sexual violence are a recognised in law as a healthcare priority in terms of both medical treatment and psychosocial support (Paredes Mosquera et al., 2018).

The extent of conflict-related sexual violence is of particular concern in Colombia given the exceptional duration of the conflict. It has continued for more than six decades, exposing multiple generations to cycles of violence. The Colombian armed conflict is officially recognised as beginning in 1964 with the formation of the guerilla group the Revolutionary Armed Forces of Colombia (known by their Spanish acronym, the FARC, later FARC-EP), following a period of bipartisan violence known as *La Violencia* (1948-1958). The origins of the conflict lie in rural inequality, political exclusion, and lack of protection from violence but its expansion and persistence are closely linked to the illicit drug economy, which has provided both funding and strategic incentives for territorial control. The conflict has been characterised by the involvement of a complex array of actors, including numerous guerrilla groups, paramilitary forces, state military units, and criminal organisations. Civilians in the conflict have experienced systematic victimisation and prolonged insecurity as groups have vied for control of populations, resources, and strategic locations, resulting in widespread displacement and social fragmentation (Restrepo & Padilla-Medina, 2023). Following four years of negotiations, a peace agreement was signed in 2016 between the Colombian state and FARC-EP. However, violence has persisted in many regions due to the continued activity of FARC-EP dissident groups, the presence of armed groups not party to the agreement, uneven implementation of the accord, and ongoing violence motivated by the protection of illicit economies (Niño & Palma, 2023).

Sexual violence has been widely reported throughout the duration of the conflict. It remains a significant public health issue and an urgent medical priority. Survivors of sexual violence are at increased risk for a range of acute and chronic medical conditions, including sexually transmitted infections, unintended pregnancies, traumatic injuries, and poor mental health (Ba & Bhopal, 2017; Stein et al., 2025). Globally, sexual violence against civilians is endemic within armed conflict, manifesting in diverse forms and intensities across different contexts, countries, and phases of conflict (Cohen & Nordås, 2014). Consequently, understanding the dynamics of sexual violence in armed conflict is essential to public health approaches, informing prevention and treatment during conflict and in post-conflict periods (Bendavid et al., 2021).

Unfortunately, understanding armed-conflict-related sexual violence within Colombia has been difficult due to a paucity of studies that look at population data at a scale commensurate with the conflict itself, although some important earlier work has laid critical groundwork. Within the Colombian armed conflict, sexual violence has been noted to occur opportunistically (Wirtz et al., 2014), as part of broader gender-based violence endemic in society (Kreft, 2020), as well as being a systematic tool used by armed actors to achieve ‘strategic’ aims such as intimidation, demoralisation, retaliation, and territorial control (Blanco Blanco et al., 2021; Céspedes-Báez, 2010). A survey conducted by Urrego et al., (2020) found a 45.1% prevalence of sexual harassment, 16.8% prevalence of rape, and 3.9% prevalence of forced prostitution, although involved focused sampling of 1,975 women aged 15-44 in highly conflict-affected areas of Colombia, and restricted the sampling range to events occurring between 2010 and 2015. Partial statistics have been published as parts of reports identifying perpetrators or elaborating the impact of sexual violence (e.g. Observatorio de Memoria y Conflicto & Centro Nacional del Memoria Historica, 2021; San Pedro, 2010) with broad consensus that the armed-conflict-related sexual violence has disproportionately affected women and girls (Guzmán & Prieto, 2014), but there are a lack of studies that have used population-level data to characterise sexual violence covering the duration of the conflict.

Public health approaches are best informed not only by an understanding of the demographic profile of affected individuals but also by evidence on how incidence spreads geographically and evolves over time, an aspect that is likely to be particularly important for understanding the patterns of violence (Naumann et al., 2019). While existing studies have described prevalence and individual risk factors, understanding how sexual violence clusters spatially and how these patterns evolve in relation to broader conflict dynamics can help target post-violence support and inform realistic timescales for prevention.

Consequently, this study aimed to characterise the demographic profile of sexual violence victims and examine the spatial and temporal dynamics of conflict-related sexual violence in Colombia. To achieve this, we used two population scale registers of armed conflict victimisation to understand the nature of sexual violence in the Colombian armed conflict: i) the national Register of Victims (RUV) – a statutory register of over 8 million people who have been affected by the armed conflict where registration is a necessary step for qualifying for reparations; and, ii) the National Centre for Historical Memory (NCMH) database of Colombian armed conflict victimisations events – a register of armed conflict events based on multiple information sources that aims to record events nationally and over the entire duration of the conflict.

We focused on two temporal windows: the entire conflict period (1964–2024) and the decade 2014–2024. Analysis of the full conflict period provides a long-term historical perspective on the spatial and temporal evolution of conflict-related sexual violence. The more recent decade coincides with the enactment of Law 1719 (2014) in Colombia, which specifically guarantees access to justice and comprehensive care for victims of sexual violence in the context of the armed conflict. An analysis of cases during this more recent time window is therefore particularly relevant for understanding the nature of sexual violence since there has been a statutory obligation to provide detection, treatment and support.

We used log-Gaussian Cox process (LGCP) modelling to estimate the latent geospatial intensity of sexual violence events. The latent spatial intensity reflects the expected concentration of sexual violence events across locations. Areas with higher intensity indicate where events tend to cluster, even in places with few or no reported cases, after accounting for population density and spatial patterns observed elsewhere. This identifies differences in underlying event concentration across regions and, by itself, does not indicate local event-to-event contagion. To assess whether such contagion is present, we used Hawkes process modelling to test for temporal contagion, where the occurrence of one event may increase the risk of subsequent additional incidents. By comparing Hawkes process models with and without adjustment for baseline rate, we aimed to distinguish the presence of short-term event-to-event contagion from slower-moving trends likely reflecting structural and contextual factors that drive sexual violence.

## Methods

The project received ethical approval from both the Ethics Committee of the Fundación Santa Fe de Bogotá in Colombia (ID CCEI-15951-2023) and the University College London Research Ethics Committee in the United Kingdom (Project ID: 8275/002).

## Data sources

We used two data sources to profile victims of sexual violence in the Colombian armed conflict. We describe each of these data sources below.

### National Register of Victims of the Colombian Armed Conflict

The *Registro Unico de Víctimas* (RUV), as it is known in Spanish, was established in 1985 and records individuals affected by the armed conflict, serving as a prerequisite for accessing state-provided reparations. Enrolment in this registry occurs at designated offices, such as municipal authorities or the national Victims’ Unit. As part of the registration process, individuals undergo an interview detailing the circumstances, and nature of the victimising events, and may be required to submit additional evidence. Under the legal framework, victims are defined as those who have experienced harm directly, or as immediate family members (including parents, children, or siblings) of those affected. Location is recorded in terms of municipality, the smallest administrative division of Colombia of which there are 1,104 in the country. Victims of sexual violence are eligible to register within a three-year period following the occurrence of the event. In 2024, legislation was passed acknowledging the unique barriers to reporting sexual violence, including a grace period extending until 2026 for victims whose prior registration attempts were denied solely due to the expiration of earlier statutory deadlines. This dataset was provided under agreement with the Colombian government and is not publicly available.

### National Centre for Historical Memory (NCMH) Database of Violent Events of the Internal Armed Conflict in Colombia

The National Centre for Historical Memory (*El Centro Nacional de Memoria Histórica* in Spanish) is a statutory institution in Colombia responsible for recording experiences of the victims of the armed conflict and was created as part of the country’s transitional justice efforts. It maintains a database of violent victimisation that occurs as part of the armed conflict and gathers and cross-references data from several sources, including government records (e.g. legal rulings, military reports), non-governmental organisation reporting, truth commissions, testimonies from victims and communities, media reports, and academic studies. Events are timestamped and geolocated with latitude and longitude. The database is open licensed and publicly available online.

## Exposure

Exposure to sexual violence is coded specifically in both databases and this was used as the exposure variable. For the Victims Register, sexual violence is defined in the Victims Unit assessment criteria (Unidad para las Víctimas, 2021, p. 3) as “Crimes against sexual freedom and integrity” and is defined as any act that violates the dignity, freedom or sexual integrity of a person through the use of physical, psychological or moral force that imposes a sexual behaviour against their will. The NCHM database does not itself have a specific definition of sexual violence, although the National Centre for Historical Memory define sexual violence as “Any act of a sexual nature imposed through the use of force, coercion, psychological oppression, abuse of power, or fear of violence” (Centro Nacional de Memoria Histórica, 2017).

## Statistical analysis

For RUV data, analysis was completed using *R* version 4.5.0 (R Core Team, 2025) on a Windows secure analysis platform. For the NCHM data, analysis was conducted with *R* version 4.5.0 on a Linux x86 platform. Analysis code and output is available as Jupyter notebooks on the online archive: https://github.com/ElisavetPappa/col-conflict-sv

Summary statistics were calculated to describe the sample, identify the profile of victims and understand co-occurrence with other types of exposure to the armed conflict. We also used two approaches to identify geospatial and temporal clustering of sexual violence events for the entire armed conflict (1964-2024) and, more recently, 2014-2024.

### Latent geospatial structure of conflict-related sexual violence

Due to the inclusion of timestamps and geographic coordinates for victimisation events, we used the NCHM Database to analyse the underlying geospatial structure of reported sexual violence incidents using Log-Gaussian Cox Process (LGCP) models (Brix & Diggle, 2001) . LGCP models estimate spatial variation in event intensity by accounting for latent factors that influence risk patterns which are not directly observable in the data but detectable through spatial clustering beyond what would occur by random chance. Using the *stelfi* package (Jones-Todd & van Helsdingen, 2024), we fitted log-Gaussian Cox process models. The LGCP model estimates three key parameters: the baseline log-intensity (LGCP β), representing the overall mean spatial intensity on the log scale; the spatial range, which indicates the distance over which the latent field exhibits strong spatial correlation (with smaller values suggesting tighter, more localised clustering); and the latent field standard deviation, quantifying the magnitude of unexplained spatial variation after accounting for covariates. Together, these parameters characterise both the overall intensity and the geographic structure of clustering. To account for population density, we rasterised back-projected census data (population divided by area) from the Colombian National Population and Housing Census available from the *ColOpenData* package (Otero et al., 2025) and extracted log-transformed values at mesh nodes. We compared this to a model that did not control for population density to assess the extent to which population density contributes to event clustering. The resulting intensity surfaces and latent fields were then projected to identify and visualise geographic clusters with elevated risk for sexual violence.

### Temporal contagion of sexual violence events

To investigate whether sexual violence events increase the probability of further sexual violence, we used the NCHM Database and Hawkes process modelling to test for temporal contagion (also known as event dependence or self-excitation) using Hawkes process models via the *stelfi* package (Jones-Todd & van Helsdingen, 2024). We added one day of random jitter to event times, which is necessary in Hawkes process models to prevent events recorded with the same time from artificially inflating contagion estimates, but means any contagion estimate of less than one day falls below the effective temporal resolution of the analysis. Parameters were estimated via maximum likelihood. The Hawkes process models estimated a baseline intensity (μ), an excitation parameter (α), and a decay rate (Hawkes β). We also calculated branching ratio (α/ Hawkes β) to quantify the proportion of events expected to trigger additional events, reflecting the degree of event-dependence in the process (Hardiman & Bouchaud, 2014). A value approaching one indicates a near-critical, self-reinforcing system, in which each event is likely to lead to subsequent events. To understand to what extent temporal contagion was accounted for by changes in background rate, we compared the unadjusted model to a model adjusted with a non-homogenous background rate function created by fitting splines to the incidence rate.

### Model validation

We completed model validation steps on both LGCP and Hawkes process models. To assess whether the LGCP adequately captured clustering of sexual violence events, we conducted simulation-based validation. To check that clusters were not artefactual interpretation of noise, we compared the observed pattern to spatial randomness using Ripley’s K-function. In addition, we simulated 1,000 point patterns from the fitted model and performed two posterior predictive checks. Firstly, we compared the distribution of total event counts across simulations to the observed count, to assess the extent to which the model reproduced the overall event frequency. Secondly, we computed spatial density estimates for the observed and simulated patterns, calculated means and 95% CIs of the simulated densities, and compared these with the observed spatial density surface to assess whether observed local events densities were consistent with the model’s posterior predictive uncertainty or reflected systematic under- or overestimation of spatial clustering. To assess the adequacy of the fitted Hawkes process models, we performed a posterior predictive check to assess whether the model reproduced timing patterns seem in the observed data. For each model, we simulated 1,000 datasets using the model parameters over the observed time window. We then compared the observed mean inter-event time to the distribution of simulated means to check that the observed value fell within the central 95% of the simulated distribution, indicating that the fitted model adequately reproduces the temporal spaces of events.

## Results

### Demographics of victims of sexual violence and differences between data sources

Demographics in terms of gender, ethnicity, and age of victims of sexual violence from each dataset are presented in Table 1.

**Table 1.**
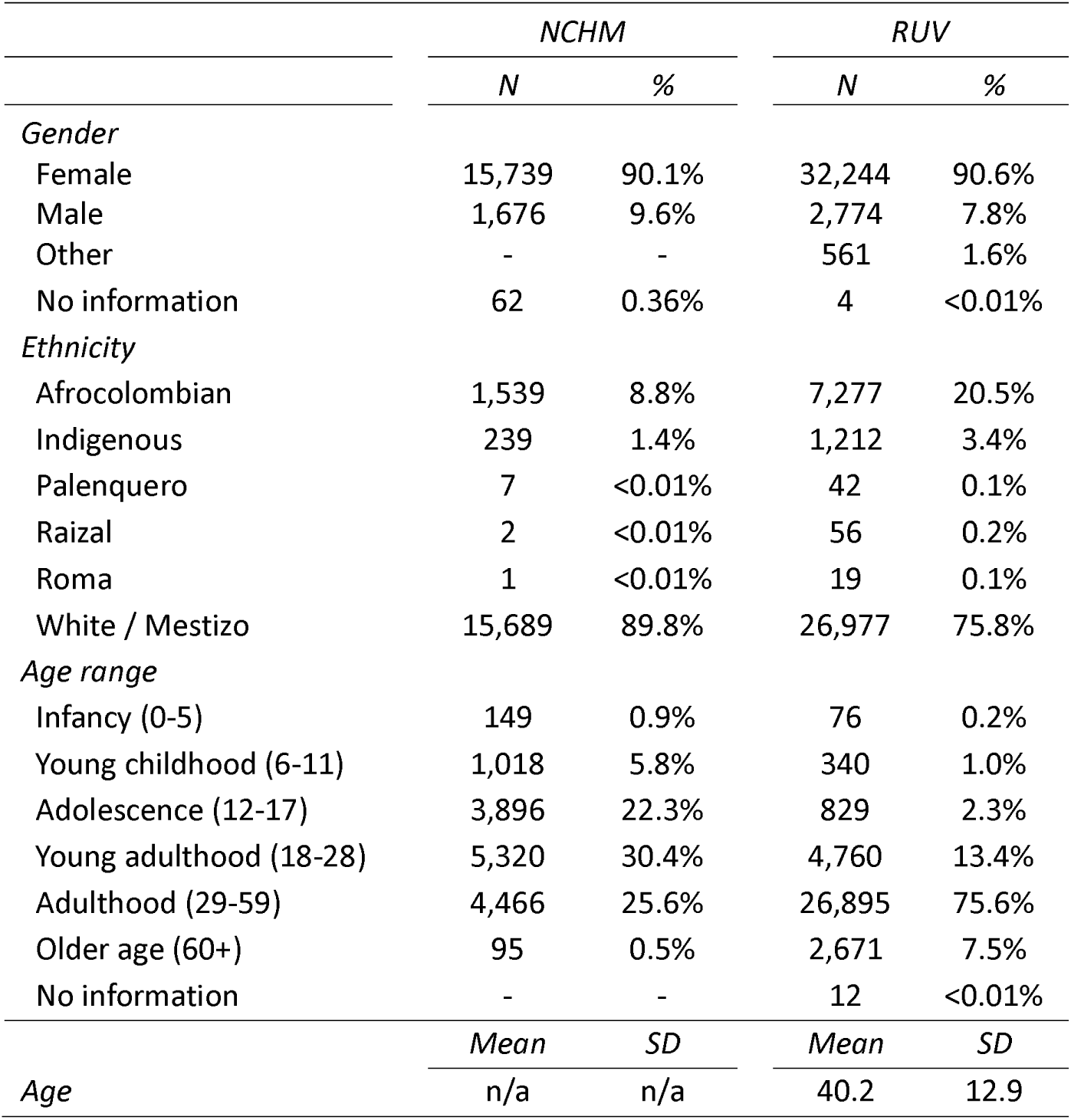
Demographics of victims of sexual violence by source of information. NCMH = National Centre for Historical Memory Database; RUV = Register of Victims Database. Age only reported by age range in NCHM.

As can be seen in Table 1, the RUV dataset has a greater proportion of ethnic minorities and an older age profile of victims than in the NCHM dataset. Perpetrators of sexual violence in terms of armed group were only recorded in the NCHM dataset but this variable had very high rates of missing data meaning it is unlikely to provide reliable information: 17,405 cases (99.59%) had no information in perpetrator, state agents were identified as perpetrators in 41 cases (0.23%), guerrilla groups in 26 cases (0.15%), paramilitary groups in 5 cases (0.03%).

### Co-occurrence with other exposures to the armed conflict

There are very few instances of co-occurrence of sexual violence events with other armed conflict exposures events in the NCHM (Table 2). Registration of sexual violence victimisation co-occurs with registration of other armed conflict exposures at much higher levels in in the Victims Register (Table 3). This data indicates that sexual violence tends to be associated with more personally directed forms of armed conflict exposure, such as torture and physical injury, rather than with property damage or group-level events such as antipersonnel mines and displacement.

**Table 2.** Co-occurrence between other Colombian armed conflict exposure events and conflict-related sexual violence in the National Centre for Historical Memory (NCHM) Database.

| *Armed conflict exposure event* | *N* | *N Co-occurrence with sexual violence* |
| --- | --- | --- |
| Witness to combat operations | 51,830 | 0 |
| Massacres | 27,580 | 12 |
| Terrorist attacks | 820 | 0 |
| Attacks on civilian property | 1,469 | 0 |
| Kidnapping | 38,580 | 4 |
| Damage to civilian property | 452 | 0 |
| Antipersonnel mines | 10,440 | 0 |
| Selective killings | 184,031 | 4 |
| Forced disappearance | 80,866 | 7 |
| Child recruitment to armed groups | 18,175 | 5 |

**Table 3.** Co-occurrence between other armed conflict exposure events and conflict-related sexual violence in the Victims Register (RUV) Database.

| *Armed conflict exposure event* | *N* | *N Co-occurrence with sexual violence* | *%* |
| --- | --- | --- | --- |
| Torture | 9,912 | 1,818 | 18.3% |
| Physical wounds | 13,880 | 1,657 | 11.9% |
| Child recruitment to armed groups | 7,756 | 455 | 5.9% |
| Kidnapping | 33,487 | 1,780 | 5.3% |
| Witness to combat | 82,845 | 1,924 | 2.3% |
| Threats | 520,805 | 9,237 | 1.8% |
| Psychological wounds | 13,949 | 226 | 1.6% |
| Loss of property | 120,168 | 996 | 0.8% |
| Forced disappearance | 160,499 | 759 | 0.5% |
| Dispossession of land | 31,464 | 160 | 0.5% |
| Forced displacement | 7,540,257 | 29,306 | 0.4% |
| Murder | 919,051 | 2,473 | 0.3% |
| Forced confinement | 75,961 | 192 | 0.3% |
| Antipersonnel mines | 11,250 | 12 | 0.1% |

### Raw geographic dispersion of sexual violence events

Although location is coded differently in each database (event by specific latitude and longitude in the NCHM Database, registration by municipality in the RUV National Victims Register), the two datasets show comparable patterns in terms of areas of high prevalence, as shown in Figure 1. In particular, higher concentrations of reported sexual violence are apparent in the Andean region and in several historically conflict-affected areas, including close to the Pacific coast, particularly Chocó, Valle del Cauca, Cauca, and Nariño, and northwestern departments such as Antioquia and Córdoba.

**Figure 1.**
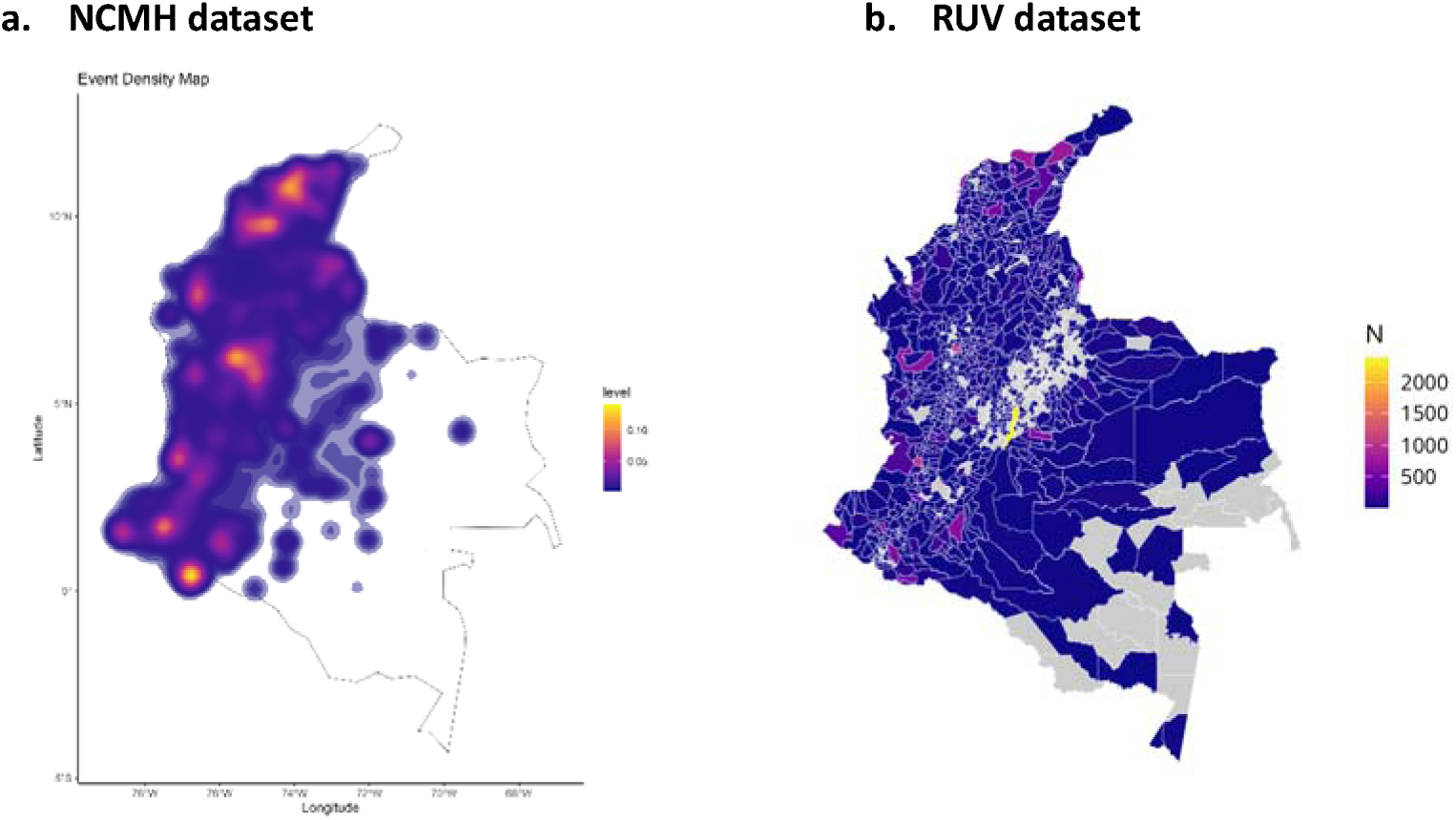
Raw density of total sexual violence events 1964-2024 as recorded in a) National Centre for Historical Memory (NCMH) database, and b) National Victims Register (RUV) by municipio in the Colombia armed conflict. Grey areas indicate no data.

### Latent geospatial structure of conflict-related sexual violence

Spatial analysis using the LGCP revealed a latent geospatial structure of sexual violence that was clustered within defined geographic extents (Figure 2), distinct from raw geospatial density (Figure 1). The broader intensity surface estimated by the LGCP model suggests that over the entire conflict period sexual violence may be more widely present than is apparent from raw event counts, and this particularly notable in areas East of the Andean region. Mesh-based integration weights are shown in Figure S1 of the supplementary material and spatial projections of the latent Gaussian Markov random fields are shown in Figure S2 of the supplementary material.

**Figure 2.**
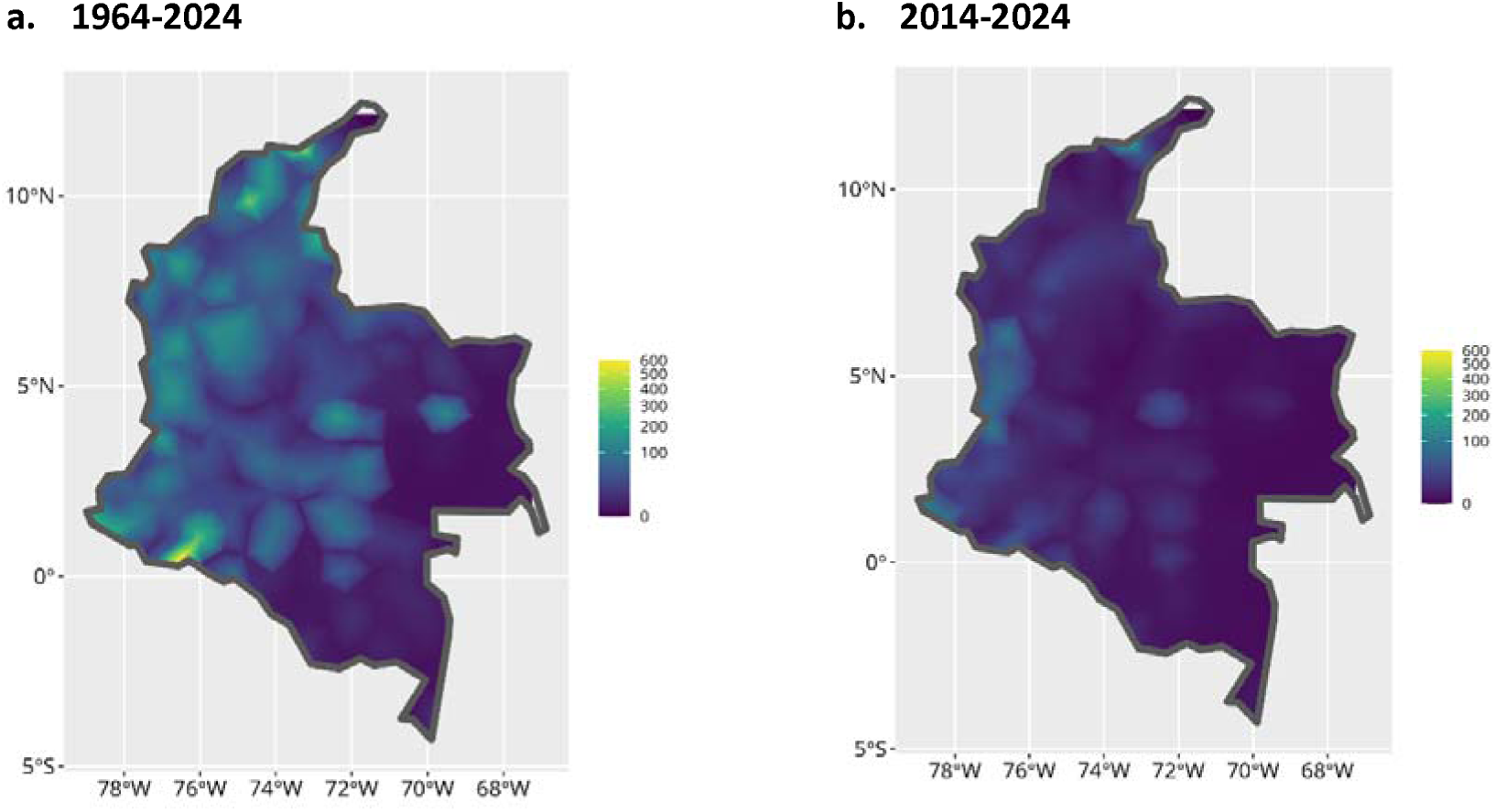
LGCP intensity plot of latent geospatial structure identifying areas of elevated probability of sexual violence for: a) entire duration of the conflict b) 2014-2024. Both adjusted for population density

For data spanning the entire conflict period (1964-2024), the unadjusted LGCP model estimated significant spatial clustering (LGCP β = 2.580; SE = 0.600) with a spatial range of 2.385 km (SE = 0.297; 95% CIs 1.802 – 2.968), indicating that locations within this distance are spatially correlated and have similar underlying event probability, and latent field standard deviation of 2.323 (SE = 0.215). After adjusting for population density, which showed a significant positive association with sexual violence events (LGCP β = 0.530; SE = 0.088), the spatial range reduced to 1.733 km (SE = 0.278), and spatial variation declined to 1.783 (SE = 0.152; 95% CIs 1.188 – 2.278), indicating that population density explained approximately 27% of the spatial range and 23% of unexplained spatial heterogeneity. For sexual violence from 2014-2024, there was a strongly reduced baseline intensity (LGCP β = 1.171; SE = 0.570 in the unadjusted model; LGCP β = 0.551, SE = 0.385 in the adjusted model). Spatial clustering extended over a wider area: 2.523 km in the unadjusted model (SE = 0.371; 95% CIs 1.797 – 3.250) vs 1.868 km (SE = 0.221; 95% CIs 1.219 – 2.517) when adjusting for population density. The population density effect remained significant in the recent period (LGCP β = 0.458; SE = 0.082), demonstrating the persistent influence of population distribution on the spatial patterning of conflict-related sexual violence throughout the conflict timeline. Full parameters for all LGCP models are shown in Tables S1 and S2 of the supplementary material. Plots for models not adjusted by population density are shown in Figures S3a and S3b of the supplementary material, alongside spatial uncertainty plots for all models (Figures S4a-b).

The LGCP model validation checks indicated adequate model fit for both time periods. Ripley’s K-function confirmed that the observed spatial distribution of sexual violence events was markedly distinct from spatial randomness. The observed event counts fell near the centre of the distribution of simulated counts from the 1,000 realisations drawn from the fitted intensity surfaces, indicating that the models accurately reproduced the expected number of events, although with a slight under-estimation of density of sexual violence events for the 1964-2024 period. Posterior predictive checks indicated that the mean simulated density surfaces closely matched the observed spatial density, reproducing the same major high intensity locations across Colombia, with the observed density falling within the 95% CIs. Simulation results are reported in Figures S5-S6 of the supplementary material.

### Temporal contagion of sexual violence events

The frequency of NCHM reported sexual violence events by year from is shown in Figure 3.

**Figure 3.**
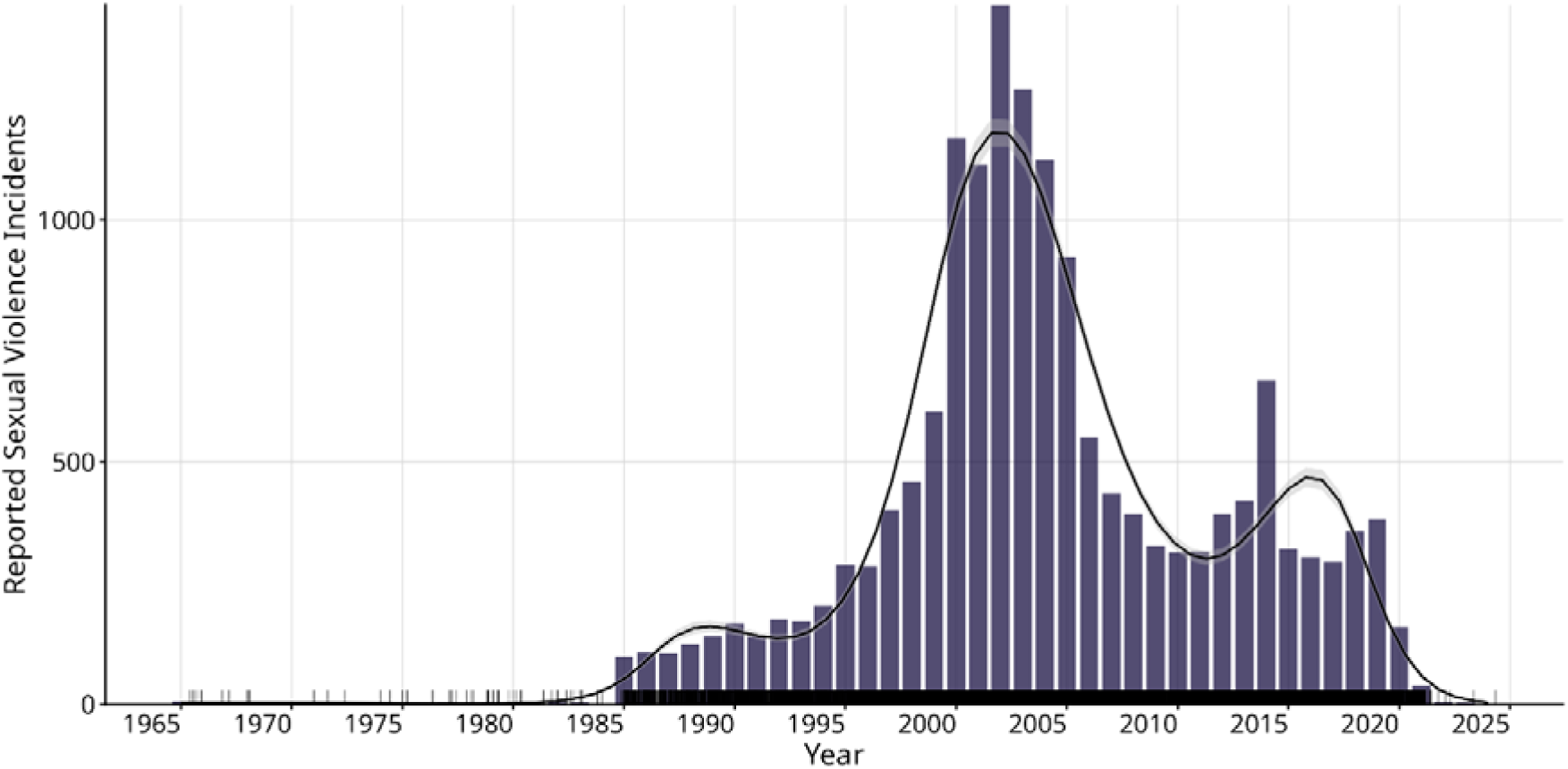
Sexual violence events reported in the National Centre for Historical Memory (NCHM) database with trend line.

Hawkes process modelling of the entire conflict data (1964-2024) indicated moderate temporal contagion of sexual violence. The Hawkes process with a constant baseline showed a low baseline rate of events (μ = 0.009; SE = 0.001), corresponding to a 0.9% chance of occurrence per day, with modest temporal dependence (α = 0.186; SE = 0.011). The decay rate (Hawkes β = 0.188; SE = 0.011) suggested that the contagion effect was time-limited, with a half-life of approximately 3.7 days (ln(2)/Hawkes β). The branching ratio (α/Hawkes β = 0.989) indicates a near-critical process, where each event generates, on average, just under one additional event. When adjusting for non-homogeneous baseline rate, there was a stronger immediate increase in intensity for subsequent events (α = 1.283; SE = 0.040) but the decay rate (Hawkes β = 3.757; SE = 0.117; half-life = 0.18 days) was below the 1-day temporal resolution of the data due to the 1-day artificial jitter added to ensure no simultaneous events as the Hawkes model assumes continuity, meaning any temporal contagion effect occurred over less than a day, suggesting negligible temporal contagion when background rate changes are accounted for. For Hawkes process modelling of sexual violence events from 2014-2024, the constant baseline model showed a lower baseline rate (μ = 0.007; SE = 0.003), corresponding to a 0.7% daily chance of occurrence, with lower temporal contagion (α = 0.069; SE = 0.010) and a longer decay rate (Hawkes β = 0.069; SE = 0.010; half-life = 10.04 days). The branching ratio (α/β = 1.000) indicated near-critical dynamics, where each event increases the expected rate of subsequent events by approximately one additional event. The model adjusted for changing baseline rates revealed higher immediate intensity increases (α = 0.537; SE = 0.068) but a decay rate below the 1-day temporal resolution of the data (Hawkes β = 2.843; SE = 0.367; half-life = 0.24 days), suggesting negligible temporal contagion when background rate changes are accounted for. Comparison between conflict periods reveals a weakening of temporal contagion over time, with sexual violence events in recent years appearing less clustered and more independent once broader fluctuations in conflict intensity are accounted for. Plots for the Hawkes process models are reported in Figures S7 and S9 in the supplementary material.

The Hawkes model validation using posterior predictive checks supported the adequacy of the Hawkes process models for the 1964-2024 and 2014-2024 time periods. For the constant baseline models, the observed mean inter-event time of 1.32 days fell within the simulated 95% interval (0.49 – 4.70 days) for 1964-2024, and 1.49 days fell within the simulated 95% interval (0.42 – 31.87 days) for 2014-2024. For the models adjusted for baseline rate of sexual violence, the observed mean inter-event time fell within the simulated 95% interval (1.26 – 1.34 days) for 1964-2024, and within the simulated 95% interval (1.23 – 1.55 days) for 2014-2024. Simulation results are reported in Figure S8 and Figure S10 of the supplementary material and goodness-of-fit metrics and reported in Figure S11 of the supplementary material.

## Discussion

We report a population-scale profile of sexual violence within the Colombian armed conflict using complete data from the statutory Register of Victims (RUV) and the National Centre for Historical Memory (NCHM) database of armed conflict victimisation. Victims of sexual violence were overwhelmingly female and disproportionally from minority ethnic groups. Sexual violence was more likely to co-occur with types of armed-conflict-related violence which more directly targeted individuals. Geospatial analysis revealed that sexual violence may be more widely present than is apparent from raw event counts and exhibited significant geographic clustering with distinct temporal patterns across conflict periods. For the entire conflict period, event density was highly clustered within relatively confined geographic ranges, with locations within approximately 2.4 km of one another sharing similar underlying probability of events. In contrast, during 2014-2024 there was reduced spatial intensity and broader geographic clustering. Analysis of temporal contagion of events indicated that event dependence was largely driven by changes in background rate because, when background rate was adjusted for, the event-to-event temporal contagion was minimal – decaying in less than 1 day, indicating that structural and contextual influences rather than incident-specific characteristics drive temporal contagion. These findings suggest that sexual violence patterns in the Colombian armed conflict have become geographically more dispersed in recent years with reduced temporal contagion effects, indicating potential changes in conflict dynamics and perpetration over time.

Research on the perpetration of sexual violence by armed groups indicate that it is not an inevitability of armed conflict but subject to group norms, leadership, conflict dynamics, and broader political conditions (Nordås & López, 2025). These may have changed significantly in recent years. Since the height of the Colombian armed conflict, the reduction in violence has been accompanied by a fragmentation of armed groups. There is now less concentrated areas of control with influence spread more thinly across wider territories (Albarracín et al., 2023; Tkacova et al., 2023). This may explain the transition from higher-intensity, geographically focused patterns of sexual violence, potentially more associated with it occurring as part of combat operations, to broader clustering with less intense and shorter-lived temporal contagion, potentially reflecting its relationship with longer-term territorial control and community subjugation, as well as a reaction to public recognition and condemnation (Blanco Blanco et al., 2021; Fiscó, 2005).

Here, geographic clusters of armed-conflict-related sexual violence indicate priority areas for provision of services for victims. Clinical response to sexual violence requires emergency and post-acute care (Alexander & Miller, 2022) as well as long-term support services (Golding, 1999; Santaularia et al., 2014). Barriers relating to stigma, fear and health service preparedness may also delay service engagement (Zinzow et al., 2022) and we note that initiatives to promote access to existing services are themselves likely to lead to increase in the number of successfully supported victims. The geographic clustering of sexual violence covers large cities, where health services are concentrated in Colombia, as well as rural areas and small towns, including those where control is disputed between armed actors and health services may be limited. This makes the equitable provision of healthcare a significant challenge due to problems with access, resources, and active interference by armed groups that particularly affect services related to women’s health and sexual violence (Lilja et al., 2024; Ramos Jaraba et al., 2020). Consequently, while specialist sexual violence pathways may be feasible in areas of high concentration of health services, better integration of sexual violence treatment and support pathways in primary and community care may be an alternative option for rural areas (Bress et al., 2019).

Colombia’s national PAPSIVI programme (an acronym from its Spanish language title *Programa de Atención Psicosocial y Salud Integral a Víctimas*, in English “Psychosocial Care and Comprehensive Health Program for Victims”) provides healthcare and psychosocial support to victims of the armed conflict nationally. Analysis of service data indicates it successfully provides priority access with appropriate support provision to victims of the conflict registered as experiencing sexual violence (Constable Fernandez, Acosta-Ortiz, García Durán, Pappa, et al., 2025; Constable Fernandez, Acosta-Ortiz, García Durán, Saunders, et al., 2025) although clinical and social outcomes, and the extent to which victims of sexual violence who have not registered as such, are being successfully supported, is currently unknown.

By comparing Hawkes process models with and without baseline adjustment, we identified that temporal contagion is largely driven by slower-moving trends in sexual violence likely driven by structural factors associated with broader conflict dynamics, rather than short-term event-to-event contagion more associated with incident-specific characteristics. Immediate intervention to prevent further incidents of sexual violence may be practical in some situations. However, these results suggest that depending solely on a policy of reactive intervention is unlikely to be the most successful strategy for prevention of armed-conflict-related sexual violence in the Colombian context because the temporal window for preventing local contagion effects may be very small – estimated at below 1 day in our study. To address these, social and structural interventions focused on prevention may be more appropriate. Indeed, interventions to reduce sexual violence by armed groups in conflict settings have focused on exactly these processes – including reducing potential victims’ exposure to situations that would allowed opportunistic violence, increasing women’s autonomy and reducing economic dependence, reducing impunity and the perception of impunity through improved legal accountability, while deploying public campaigns aimed at increasing awareness and reducing acceptance of sexual violence (Spangaro et al., 2013). More recent initiatives have developed and deployed specific behaviour change interventions for both state and non-state armed actors aimed at reducing the perpetration of armed conflict-related sexual violence (ICRC, 2025), including in Colombia (ICRC, 2024), although the effectiveness of these interventions has yet to be determined.

We note some limitations of this study. With regard to the demographic data, the profile of victims is relatively consistent across data sources with some notable differences. Victims in the National Register of Victims were older than victims in the NCHM database, likely due to the fact that while the NCHM database records specific events, the National Register of Victims is a record of victims’ registrations, who may register several years after experiencing sexual violence. There is a higher proportion of victims from ethnic minorities in the Register of Victims, potentially reflecting its sole reliance on self-registration rather than a third-party reports, the latter of which may be more limited in areas with large populations of primarily Indigenous and Afrocolombian populations.

We also note that the diversity of sexual violence may not be well accounted for by the single coding of ‘sexual violence’ used in this study. Sexual violence may variously include rape, forced prostitution, sexual slavery, forced abortion, forced nudity, or sexual assault, to name but a few relevant criminal acts (Koos, 2017) and it is likely there may be different geographical spread and dynamics that depend on the specifics of these acts.

In addition, conflict-related sexual violence is likely under-reported and under-detected with good evidence that this is the case in Colombia (Otero & Melo, 2017). Nevertheless, different national data sources, including crime statistics, armed conflict victim registrations, and multi-source identification, tend to show similar relative changes in incidence in sexual violence that track changes in the intensity of the conflict indicating they may be sampling a similar population (Castillejo Cuéllar et al., 2022). Nonetheless, the extent to which there are specific groups who are more likely to have incidents of sexual violence remain unrecorded is not yet fully understood. Indeed, we note that individuals living in areas of active conflict (Bahamón et al., 2009), displaced women (Wirtz et al., 2014), sexual and gender minorities (Díaz Botia, 2014), and people from ethnic minorities (Marciales Montenegro, 2015) have been identified as facing particular structural barriers in reporting sexual violence and accessing care.

However, there is not a simple relationship between under-reporting and under-recognition in the geospatial models reported in this study. Log-Gaussian Cox process modelling is able to identify areas of likely high prevalence despite low reporting based on inferences from spatial clustering patterns and population density. Nevertheless, selective under-recognition may occur where barriers to reporting are unevenly geographically distributed in a way that is not accounted for differences in population density. For example, where populations of similar densities and similar underlying exposure to sexual violence show different levels of reporting due to differences in stigma, chance of reprisals, or accessibility of services. Although differences in reported and inferred prevalence may indicate areas at higher risk of undetected sexual violence, under-reporting is a widely-recognised feature of sexual violence and addressing this challenge requires dedicated initiatives. Although this issue remains under-studied in Colombia, evidence from a range of conflicts suggests that sexual violence reporting and detection may be improved by destigmatising victims; reducing risks faced by survivors; strengthening trust in institutions; ensuring the availability of multiple, accessible reporting and detection pathways; systematically curating data; guaranteeing that reports lead to meaningful responses; and implementing gender- and culture-sensitive services (Bastick & Ghittoni, 2024; Davies & True, 2018; Spangaro et al., 2015; Stark et al., 2017) and these are likely important initiatives to apply in Colombia.

In conclusion, the trajectory of sexual violence in the Colombian armed conflict reflects a shift from geographically concentrated, high-intensity incidents toward more diffuse patterns, with clear areas of high historical prevalence where victim support remains essential. The temporal dynamics of sexual violence suggest that interventions that proactively address structural and long-term contextual determinants may be an important component of prevention.

## Data Availability

The NCMH dataset used in the LGCP and Hawkes process model analyses is the open dataset from the Centro Nacional de Memoria Historica which can be accessed via https://github.com/ElisavetPappa/col-conflict-sv The RUV dataset is not available publicly.

## Acknowledgements

Many thanks to our advisors with lived experience of the Colombian armed conflict who suggested a focus on better understanding sexual violence and who inspired this study.

## Data availability statement

We used two data sources in this study. National Register of Victims of the Colombian Armed Conflict was provided under agreement with the Colombian government and is not publicly available. The National Centre for Historical Memory dataset is open data and available on their webpage: https://micrositios.centrodememoriahistorica.gov.co/observatorio/portal-de-datos/base-de-datos/ The full dataset we used in this study is available on the open GithHub archive: https://github.com/ElisavetPappa/col-conflict-sv

## References

1. Albarracín, J., Corredor-Garcia, J., Milanese, J. P., Valencia, I. H., & Wolff, J. (2023). Pathways of post-conflict violence in Colombia. Small Wars & Insurgencies, 34(1), 138–164. 10.1080/09592318.2022.2114244

2. Alexander, K. A., & Miller, E. (2022). Sexual Violence—Another Public Health Emergency. JAMA Network Open, 5(10), e2236285. 10.1001/jamanetworkopen.2022.36285

3. Ba, I., & Bhopal, R. S. (2017). Physical, mental and social consequences in civilians who have experienced war-related sexual violence: A systematic review (1981–2014). Public Health, 142, 121–135. 10.1016/j.puhe.2016.07.019

4. Bahamón, S. O., Márquez, V. Q., & Bolívar, I. (2009). Las barreras invisibles del registro de la violencia sexual en el conflicto armado colombiano. Revista Forensis, 335–349.

5. Bastick, M., & Ghittoni, M. (2024). Security Sector Reform and Conflict-Related Sexual Violence. DCAF – Geneva Centre for Security Sector Governance.

6. Bendavid, E., Boerma, T., Akseer, N., Langer, A., Malembaka, E. B., Okiro, E. A., Wise, P. H., Heft-Neal, S., Black, R. E., Bhutta, Z. A., Bhutta, Z., Black, R., Blanchet, K., Boerma, T., Gaffey, M., Langer, A., Spiegel, P., Waldman, R., & Wise, P. (2021). The effects of armed conflict on the health of women and children. The Lancet, 397(10273), 522–532. 10.1016/S0140-6736(21)00131-8

7. Blanco Blanco, J., Téllez Navarro, R. F., Bocanegra Acosta, H., Blanco Blanco, J., Téllez Navarro, R. F., & Bocanegra Acosta, H. (2021). La violencia sexual contra la mujer como el proceder táctico de los grupos armados ilegales en el marco del conflicto armado interno colombiano: Los problemas de la visibilización, la prevención y la atención. Revista republicana, (30), 125–146. 10.21017/rev.repub.2021.v30.a99

8. Bress, J., Kashemwa, G., Amisi, C., Armas, J., McWhorter, C., Ruel, T., Ammann, A. J., Mukwege, D., & Butler, L. M. (2019). Delivering integrated care after sexual violence in the Democratic Republic of the Congo. BMJ Global Health, 4(1). 10.1136/bmjgh-2018-001120

9. Brix, A., & Diggle, P. J. (2001). Spatiotemporal prediction for log-Gaussian Cox processes. Journal of the Royal Statistical Society: Series B (Statistical Methodology), 63(4), 823–841. 10.1111/1467-9868.00315

10. Castillejo Cuéllar, A., Franco Agudelo, S., Ganem Maloof, K., & Roux Rengifo, F. J. de. (2022). Mi cuerpo es la verdad: Experiencias de mujeres y personas LGBTIQ+ en el conflicto armado. Comisión de la Verdad de Colombia.

11. Centro Nacional de Memoria Histórica. (2017). La guerra inscrita en el cuerpo: Informe nacional sobre violencia sexual en el conflicto armado. Centro Nacional de Memoria Histórica.

12. Céspedes-Báez, L.-M. (2010). La violencia sexual en contra de las mujeres como estrategia de despojo de tierras en el conflicto armado colombiano. Estudios Socio-Jurídicos, 12(2), 273–304.

13. Cohen, D. K., & Nordås, R. (2014). Sexual violence in armed conflict: Introducing the SVAC dataset, 1989–2009. Journal of Peace Research, 51(3), 418–428. 10.1177/0022343314523028

14. Constable Fernandez, C., Acosta-Ortiz, A., García Durán, M. C., Pappa, E., Saunders, R., Solmi, F., Tamayo-Agudelo, W., Idrobo, F., & Bell, V. (2025). Intervention provision and engagement in Colombia’s PAPSIVI: A national psychosocial support service for over half a million victims of armed conflict (p. 2025.07.08.25331104). medRxiv. 10.1101/2025.07.08.25331104

15. Constable Fernandez, C., Acosta-Ortiz, A., García Durán, M. C., Saunders, R., Solmi, F., Tamayo-Agudelo, W., Idrobo, F., & Bell, V. (2025). Armed conflict exposure types are not equally associated with access to psychosocial support: A study of over 8[million victims of the Colombian armed conflict. International Journal of Social Psychiatry, 00207640251336726. 10.1177/00207640251336726

16. Davies, S. E., & True, J. (2018). The politics of counting and reporting conflict-related sexual and gender-based violence: The case of Myanmar. In The Difference that Gender Makes to International Peace and Security. Routledge.

17. Díaz Botia, Y. T. (2014). La reparación integral a víctimas LGBT de desplazamiento forzado y de abuso sexual en el contexto del conflicto armado colombiano. Justicia y Derecho, (2), 61–71.

18. Fiscó, S. (2005). Atroces realidades: La violencia sexual contra la mujer en el conflicto armado colombiano. Papel Político, 17, 119–159.

19. Golding, J. M. (1999). Sexual-Assault History and Long-Term Physical Health Problems: Evidence From Clinical and Population Epidemiology. Current Directions in Psychological Science, 8(6), 191–194. 10.1111/1467-8721.00045

20. Guzmán, D. E., & Prieto, S. C. (2014). Acceso a la justicia: Mujeres, conflicto armado y justicia. Djusticia.

21. Hardiman, S. J., & Bouchaud, J.-P. (2014). Branching-ratio approximation for the self-exciting Hawkes process. Physical Review E, 90(6), 062807. 10.1103/PhysRevE.90.062807

22. ICRC. (2024). Addressing Sexual Violence. International Committee of the Red Cross. https://www.icrc.org/sites/default/files/topic/file_plus_list/2024_specialappeal_sexual_violence_icrc.pdf

23. ICRC. (2025). Promising Pathways for the Prevention of Sexual Violence. International Committee of the Red Cross. https://www.icrc.org/sites/default/files/2025-07/Promising_Pathways_PSVP_2024.pdf

24. Jones-Todd, C. M., & van Helsdingen, A. B. M. (2024). stelfi: An R package for fitting Hawkes and log-Gaussian Cox point process models. Ecology and Evolution, 14(2), e11005. 10.1002/ece3.11005

25. Koos, C. (2017). Sexual violence in armed conflicts: Research progress and remaining gaps. Third World Quarterly, 38(9), 1935–1951. 10.1080/01436597.2017.1322461

26. Kreft, A.-K. (2020). Civil society perspectives on sexual violence in conflict: Patriarchy and war strategy in Colombia. International Affairs, 96(2), 457–478. 10.1093/ia/iiz257

27. Lilja, J., Ferrari, G., Alvarado, J., Fabich, L.-A., Kyzy, G. A., Kenny, L., & Hossain, M. (2024). Territorial control by non-state armed groups and gendered access to healthcare in conflict using a new complex adaptive systems framework. Humanities and Social Sciences Communications, 11(1), 855. 10.1057/s41599-024-03345-2

28. Lopera, M., & Concha, S. M. R. (2022). La violencia sexual en el marco del conflicto armado colombiano: Sobre su priorización en la JEP. Pensamiento Jurídico, (56), 183–227.

29. Marciales Montenegro, C. X. (2015). Violencia sexual en el conflicto armado colombiano: Racismo estructural y violencia basada en género. Revista Via Iuris, (19), 69–90.

30. Naumann, R. B., Austin, A. E., Sheble, L., & Lich, K. H. (2019). System Dynamics Applications to Injury and Violence Prevention: A Systematic Review. Current Epidemiology Reports, 6(2), 248–262. 10.1007/s40471-019-00200-w

31. Niño, C., & Palma, D. (2023). Transforming conflict and transforming violence: Determinants in the geometry of violence in Colombia. Critical Studies on Security, 11(3), 215–229. 10.1080/21624887.2023.2238999

32. Nordås, R., & López, E. (2025). Recent Advances in the Study of Conflict-Related Sexual Violence. Current Psychiatry Reports, 27(1), 66–75. 10.1007/s11920-024-01575-4

33. Observatorio de Memoria y Conflicto, & Centro Nacional del Memoria Historica. (2021). Estudio de Violencia Sexual en Colombia. https://saga.unodc.org.co/sites/default/files/webform/catalogacion_de_sentencias_judic/48/estudio-vs-omc-20211209-v2.pdf

34. Otero, J., Tavera Cifuentes, M. C., Hartgerink, C., & Gurson, H. (2025). ColOpenData [Computer software]. https://github.com/epiverse-trace/ColOpenData

35. Otero, M. A. G., & Melo, M. E. I. (2017). Detrás de las cifras de violencia contra las mujeres en Colombia. Sociedad y Economía, (32), 41–64.

36. Paredes Mosquera, H. H., Guachetá Torres, J. D., & Paredes Londoño, E. J. (2018). Las víctimas de violencia sexual en el marco del conflicto armado en relación con los procesos de paz en Colombia, 1991 a 2017. Jurídicas, 15(1), 88–109.

37. R Core Team. (2025). R: A language and environment for statistical computing (Version 4.5.0) [Computer software]. R Foundation for Statistical Computing. https://www.R-project.org/

38. Ramos Jaraba, S. M., Quiceno Toro, N., Ochoa Sierra, M., Ruiz Sánchez, L., García Jiménez, M. A., Salazar-Barrientos, M. Y., Bedoya Bedoya, E., Vélez Álvarez, G. A., Langer, A., Gausman, J., & Garcés-Palacio, I. C. (2020). Health in conflict and post-conflict settings: Reproductive, maternal and child health in Colombia. Conflict and Health, 14(1), 33. 10.1186/s13031-020-00273-1

39. Restrepo, M. T., & Padilla-Medina, D. (2023). Armed conflict exposure and mental health: Examining the role of imperceptible violence. *Medicine*, Conflict and Survival, 39(3), 199–221. 10.1080/13623699.2023.2222360

40. San Pedro, P. (2010). Sexual Violence in Colombia: Instrument of war. Oxfam.

41. Santaularia, J., Johnson, M., Hart, L., Haskett, L., Welsh, E., & Faseru, B. (2014). Relationships between sexual violence and chronic disease: A cross-sectional study. BMC Public Health, 14(1), 1286. 10.1186/1471-2458-14-1286

42. Spangaro, J., Adogu, C., Ranmuthugala, G., Davies, G. P., Steinacker, L., & Zwi, A. (2013). What Evidence Exists for Initiatives to Reduce Risk and Incidence of Sexual Violence in Armed Conflict and Other Humanitarian Crises? A Systematic Review. PLOS ONE, 8(5), e62600. 10.1371/journal.pone.0062600

43. Spangaro, J., Adogu, C., Zwi, A. B., Ranmuthugala, G., & Davies, G. P. (2015). Mechanisms underpinning interventions to reduce sexual violence in armed conflict: A realist-informed systematic review. Conflict and Health, 9(1), 19. 10.1186/s13031-015-0047-4

44. Stark, L., Sommer, M., Davis, K., Asghar, K., Baysa, A. A., Abdela, G., Tanner, S., & Falb, K. (2017). Disclosure bias for group versus individual reporting of violence amongst conflict-affected adolescent girls in DRC and Ethiopia. PLOS ONE, 12(4), e0174741. 10.1371/journal.pone.0174741

45. Stein, C., Flor, L. S., Gil, G. F., Khalil, M., Herbert, M., Aravkin, A. Y., Arrieta, A., Baeza de Robba, M. J., Bustreo, F., Cagney, J., Calderon-Anyosa, R. J. C., Carr, S., Chandan, J. K., Chandan, J. S., Coll, C. V. N., de Andrade, F. M. D., de Andrade, G. N., Debure, A. N., DeGraw, E., … Gakidou, E. (2025). The health effects associated with physical, sexual and psychological gender-based violence against men and women: A Burden of Proof study. Nature Human Behaviour, 9(6), 1201–1216. 10.1038/s41562-025-02144-2

46. Tkacova, K., Idler, A., Johnson, N., & López, E. (2023). Explaining conflict violence in terms of conflict actor dynamics. Scientific Reports, 13(1), 21187. 10.1038/s41598-023-48218-x

47. Unidad para las Víctimas. (2021). Manual Criterios de Valoración (Versión 3). https://www.unidadvictimas.gov.co/en/documentos_bibliotec/manual-criterios-de-valoracion-v3/

48. Unidad para las Víctimas. (2023). Boletín # 4 Datos Para La Paz. https://datospaz.unidadvictimas.gov.co/archivos/datosPaz/Boletin_Datos_para_la_Paz_JUNIO_V_FINAL.pdf

49. Urrego, A. C., Cruz, N. M., & Márquez, J. G. B. (2020). Aportes y asuntos críticos en la medición de la violencia sexual contra las mujeres en el marco del conflicto armado en Colombia: Una reflexión a partir del diseño y los resultados de la Envise 2010-2015. Estudios Socio-Jurídicos, 22(2), 1–32. 10.12804/revistas.urosario.edu.co/sociojuridicos/a.7891

50. Wirtz, A. L., Pham, K., Glass, N., Loochkartt, S., Kidane, T., Cuspoca, D., Rubenstein, L. S., Singh, S., & Vu, A. (2014). Gender-based violence in conflict and displacement: Qualitative findings from displaced women in Colombia. Conflict and Health, 8(1), 10. 10.1186/1752-1505-8-10

51. Zinzow, H. M., Littleton, H., Muscari, E., & Sall, K. (2022). Barriers to Formal Help-seeking following Sexual Violence: Review from within an Ecological Systems Framework. Victims & Offenders, 17(6), 893–918. 10.1080/15564886.2021.1978023

