## Supplementary material for "Understanding Sexual Violence in the Colombian Armed Conflict: Victim Characteristics, Spatial Clustering, and Temporal Contagion"

|  | *1964-2024* | |  | *2014-2024* | |
| --- | --- | --- | --- | --- | --- |
| *Parameter* | *Estimate* | *SE* |  | *Estimate* | *SE* |
| Β_0_ (intercept) | 2.032 | 0.369 |  | 0.551 | 0.385 |
| Β_1_ (covariate) | 0.530 | 0.088 |  | 0.458 | 0.082 |
| log τ | -2.334 | 0.121 |  | -2.170 | 0.134 |
| log κ | 0.490 | 0.161 |  | 0.415 | 0.177 |
| Range | 1.733 | 0.278 |  | 1.868 | 0.331 |
| Standard deviation | 1.783 | 0.152 |  | 1.632 | 0.159 |
| Expected no. of events | 16251.87 | - |  | 2529.286 | - |

**Table S1.** Parameter estimates for Log-Gaussian Cox Process (LGCP) model of sexual violence events in Colombia adjusted for population density

|  | *1964-2024* | |  | *2014-2024* | |
| --- | --- | --- | --- | --- | --- |
| *Parameter* | *Estimate* | *SE* |  | *Estimate* | *SE* |
| β (intercept) | 2.580 | 0.600 |  | 1.171 | 0.570 |
| log τ | -2.279 | 0.085 |  | -2.097 | 0.101 |
| log κ | 0.170 | 0.125 |  | 0.114 | 0.147 |
| Range | 2.385 | 0.297 |  | 2.523 | 0.371 |
| Standard deviation | 2.323 | 0.215 |  | 2.523 | 0.371 |
| Expected no. of events | 16253.06 | - |  | 2530.72 | - |

**Table S2.** Parameter estimates for Log-Gaussian Cox Process (LGCP) model of sexual violence events in Colombia without adjustment for population density

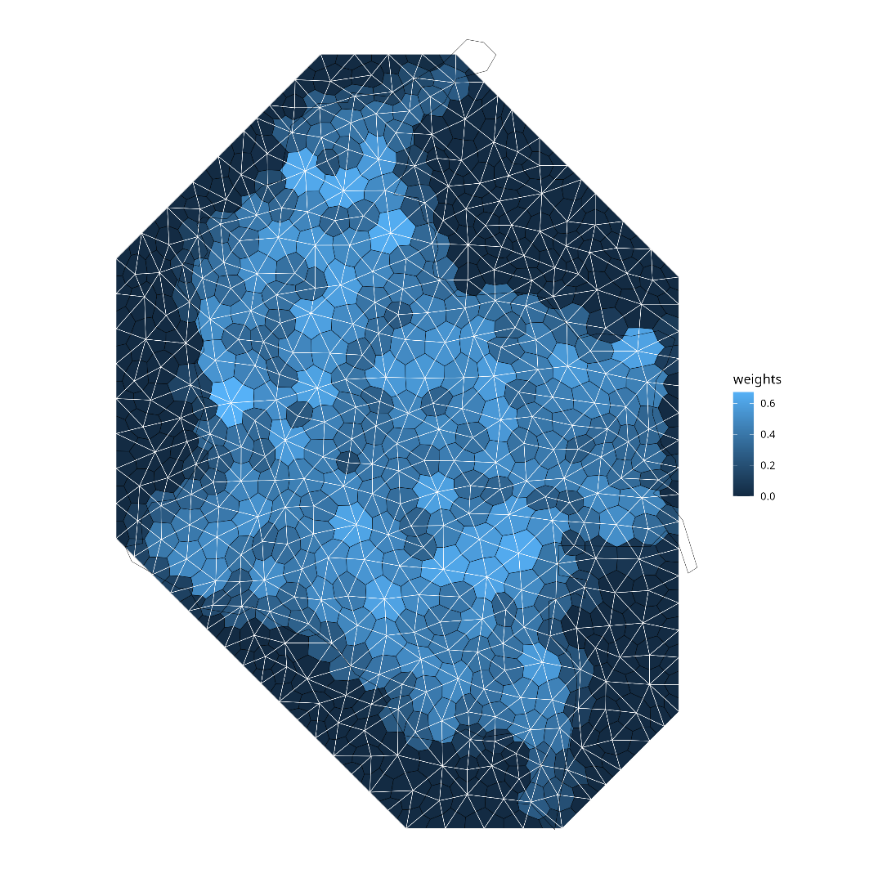

**Figure S1.** Mesh-based integration weights for log-Gaussian Cox process models

| a. **1964-2024 (population adjusted)**   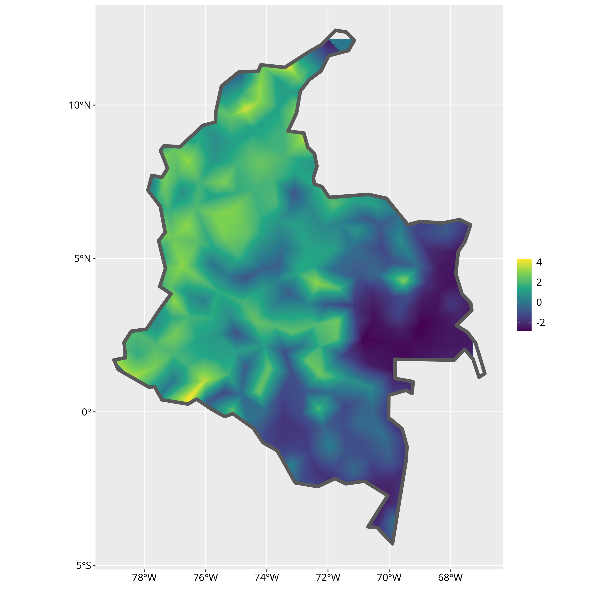 | b. **1964-2024 (unadjusted)**   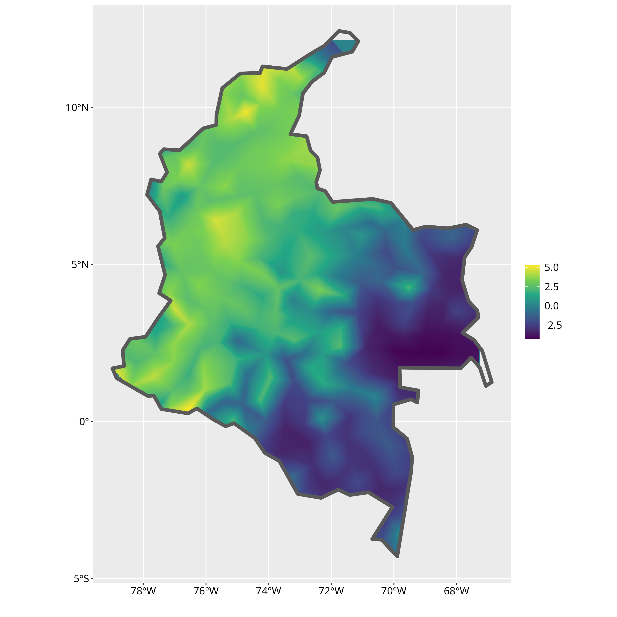 |
| --- | --- |
| c. **2014-2024 (population adjusted)**   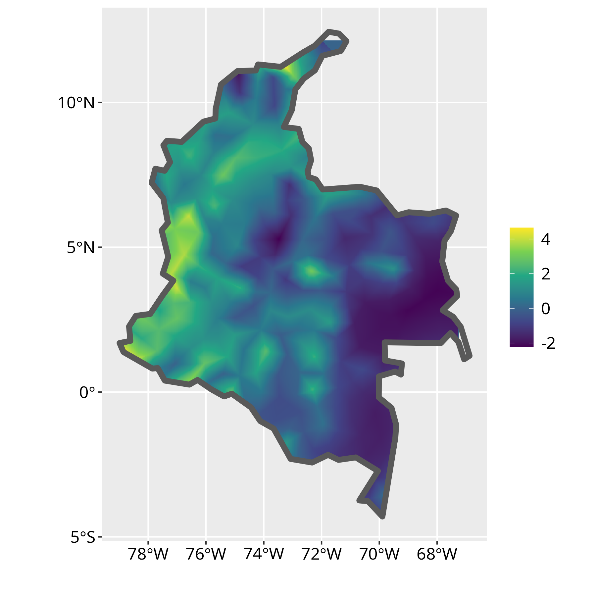 | d. **2014-2024 (unadjusted)**   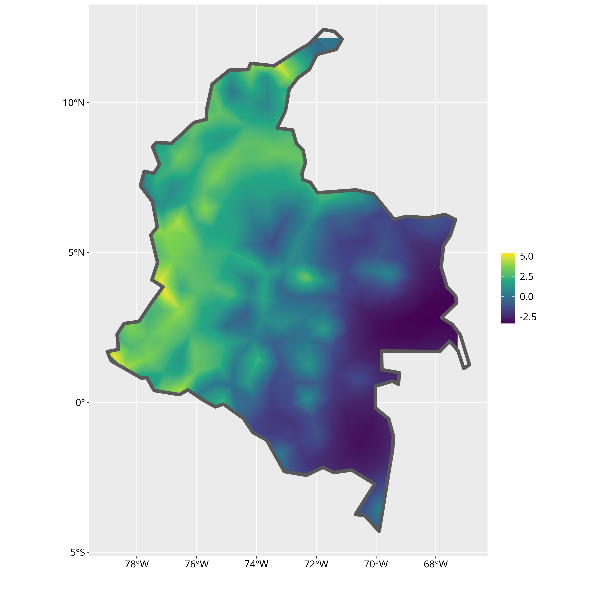 |
| **Figure S2.** Spatial projection of the latent Gaussian Markov random field (GMRF) from the fitted log-Gaussian Cox process model of sexual violence events adjusted and unadjusted for population density: | |

a. **1964-2024**

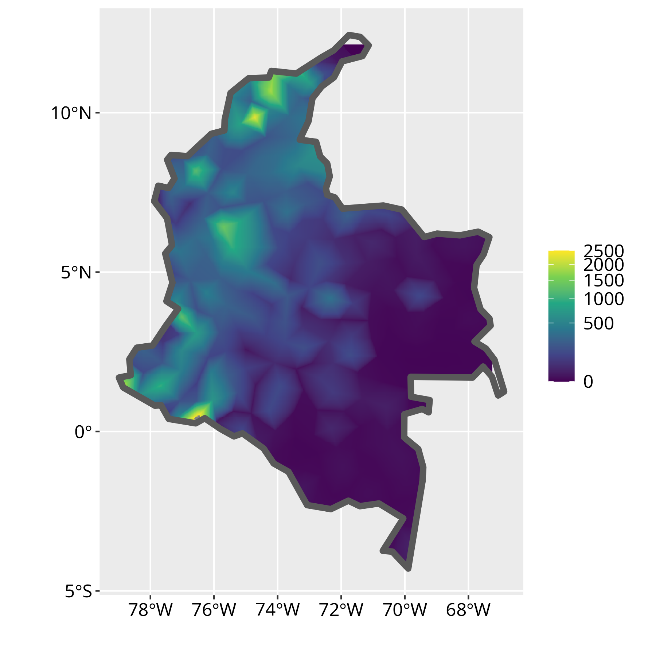

b. **2014-2024**

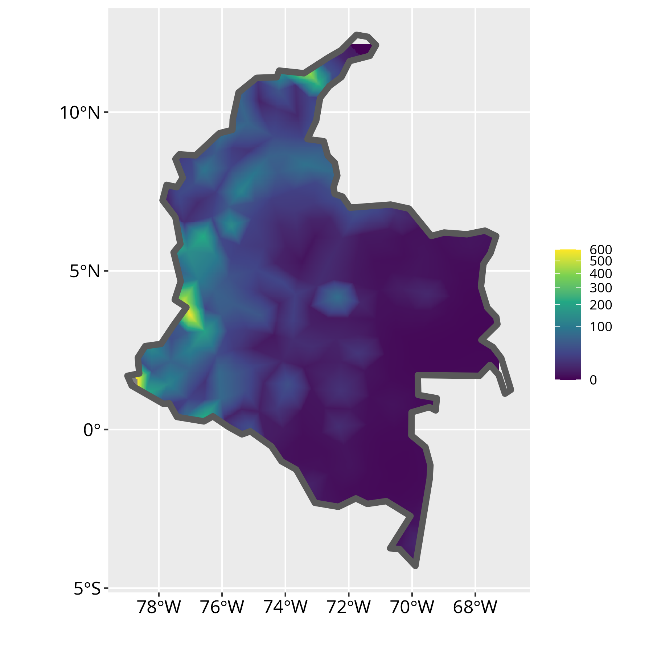

**Figure S3.** Log-Gaussian Cox Process intensity plot identifying areas of elevated risk for sexual violence for: **a)** entire duration of the conflict (1964-2024) **b)** 2014-2024. Both calculated without adjustment for population density

| a. **1964-2024 (population adjusted)**   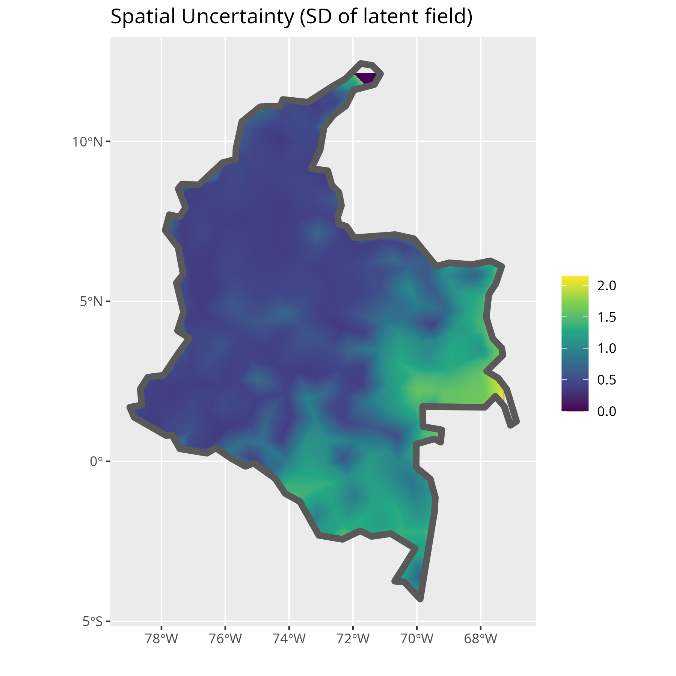 | b. **1964-2024 (unadjusted)**   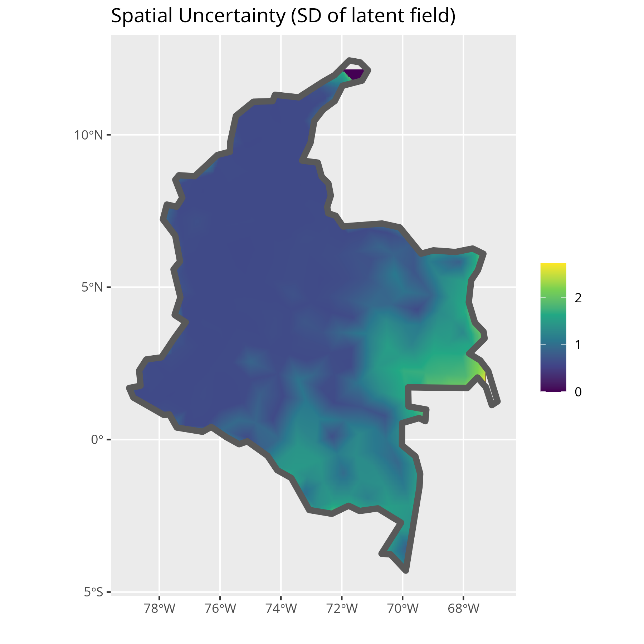 |
| --- | --- |
| c. **2014-2024 (population adjusted)**   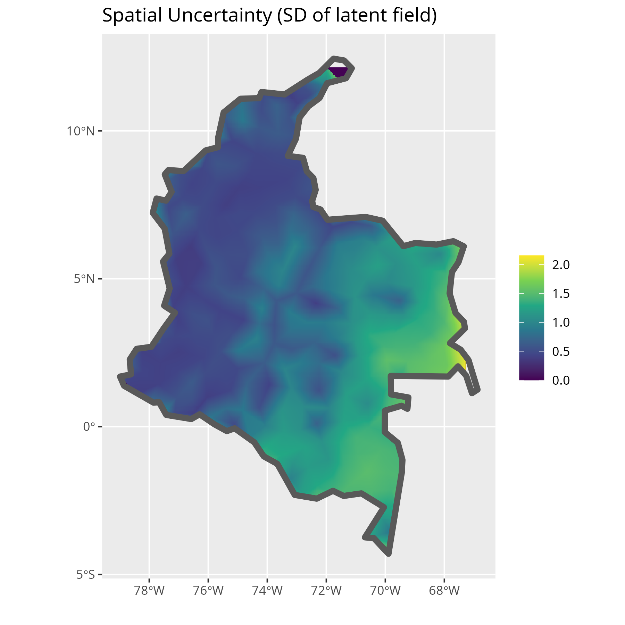 | d. **2014-2024 (unadjusted)**   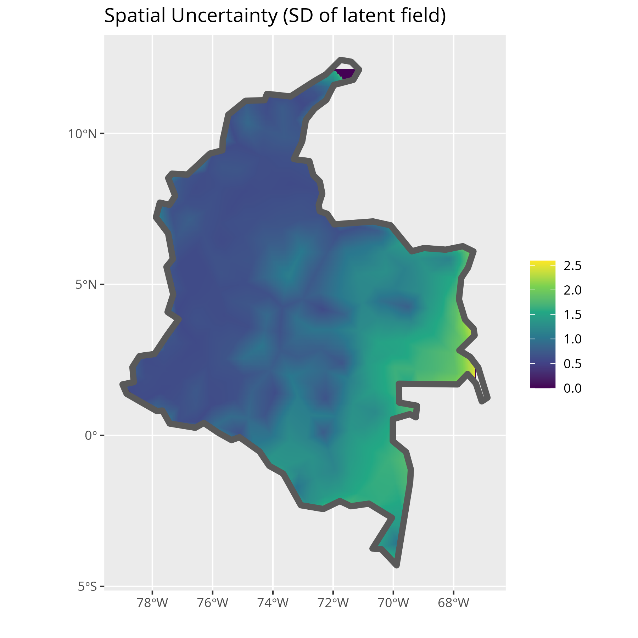 |
| **Figure S4.** Spatial uncertainty plots showing standard deviation of latent field | |

| **a)**  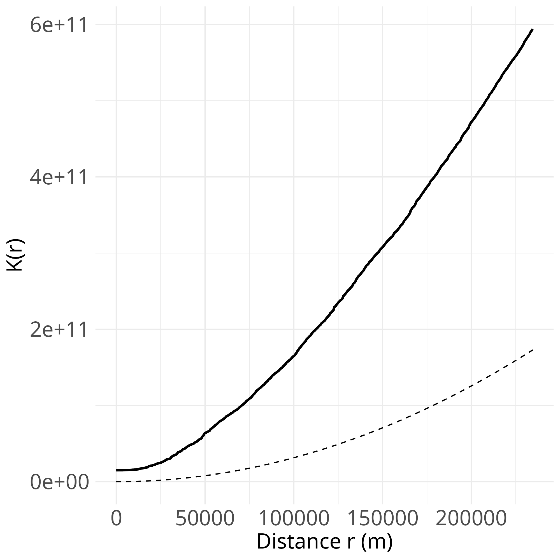 | **b)**  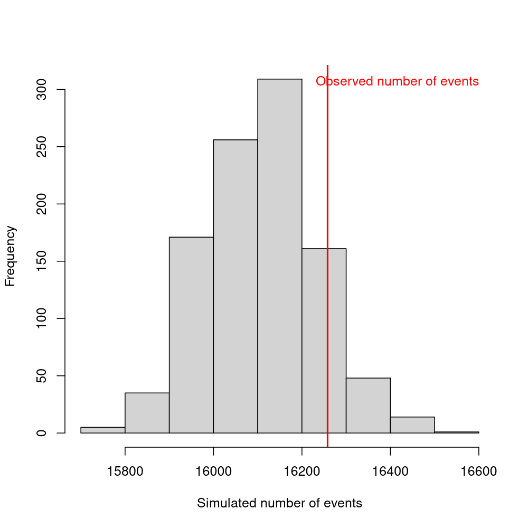 |
| --- | --- |
| **c)**  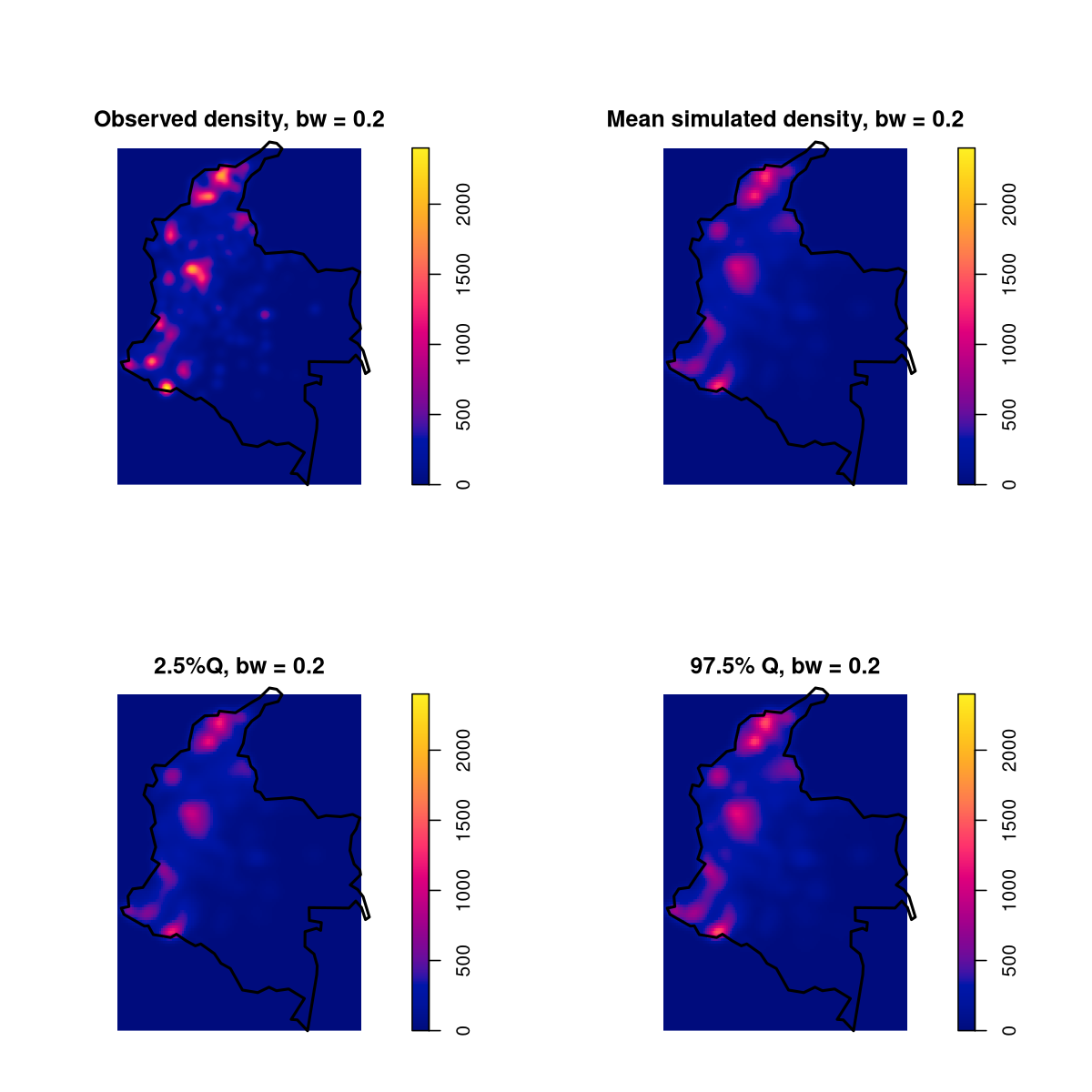 | |
| **Figure S5.** Validation test results for 1964-2024 population adjusted data. **a)** Ripley’s K-function; **b)** simulated vs observed number of events; **c)** mean and 95% CIs of simulated datasets from intensity surface in comparison to observed density | |

| **a)**  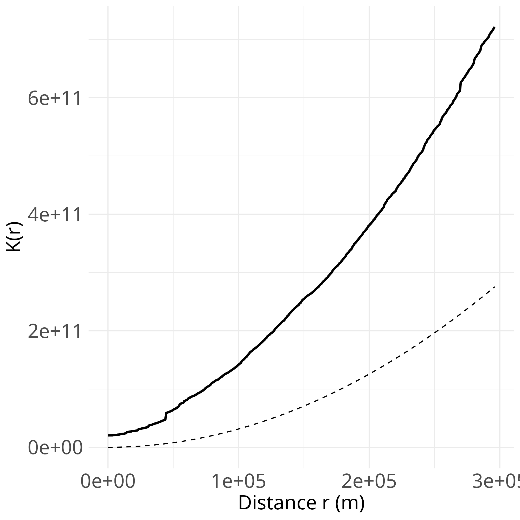 | **b)**  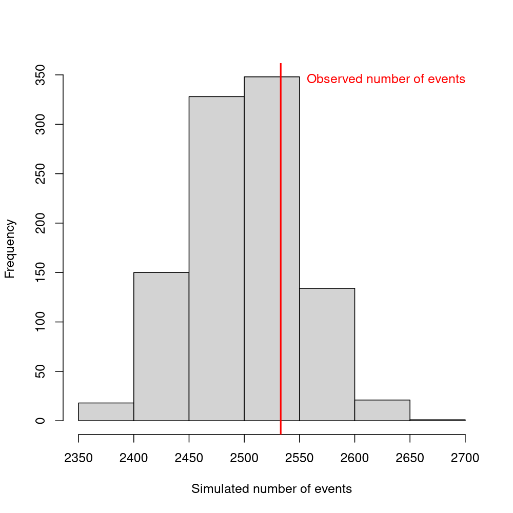 |
| --- | --- |
| **c)**  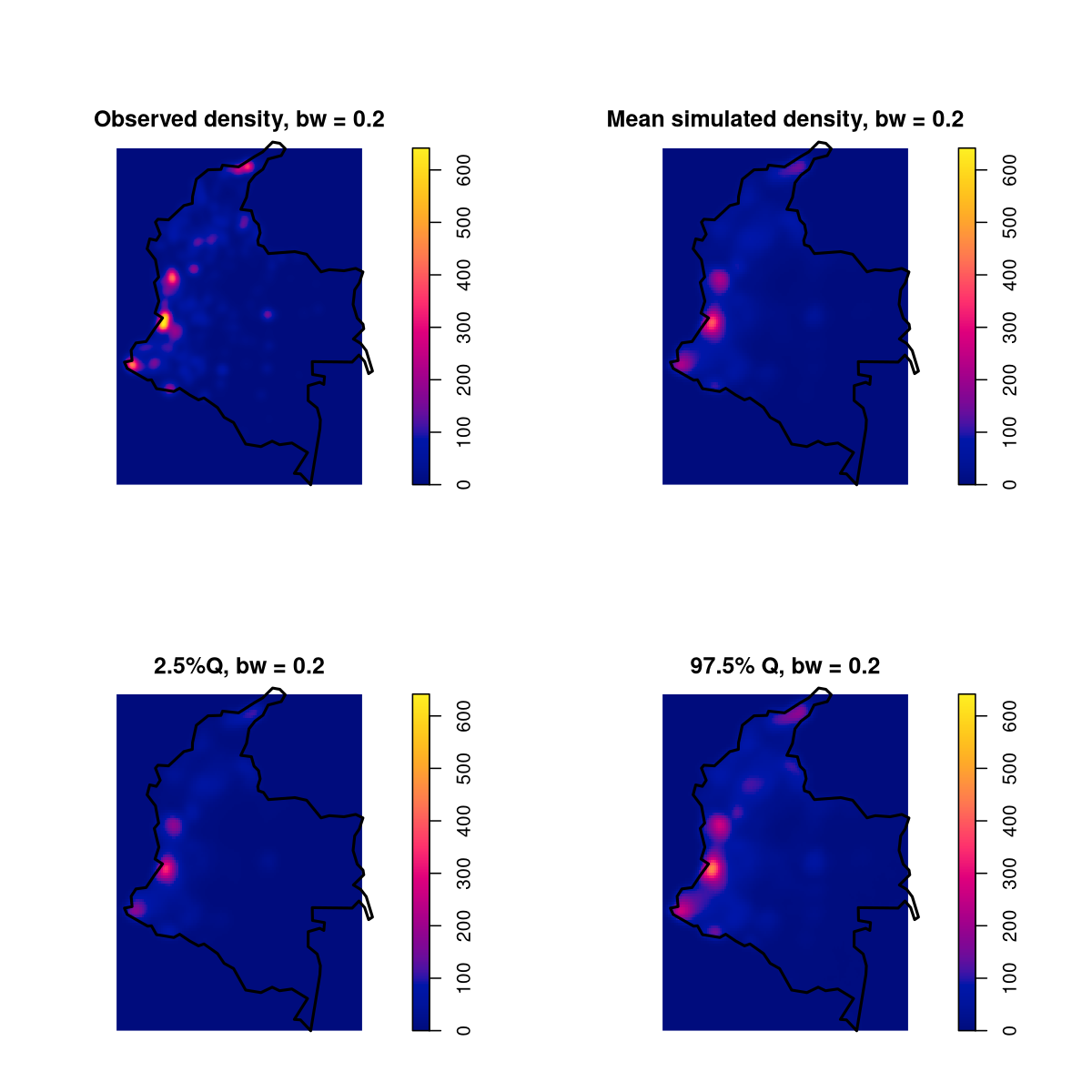 | |
| **Figure S6.** Validation test results for 2014-2024 population adjusted data. **a)** Ripley’s K-function; **b)** simulated vs observed number of events; **c)** mean and 95% CIs of simulated datasets from intensity surface in comparison to observed density | |

**a)**
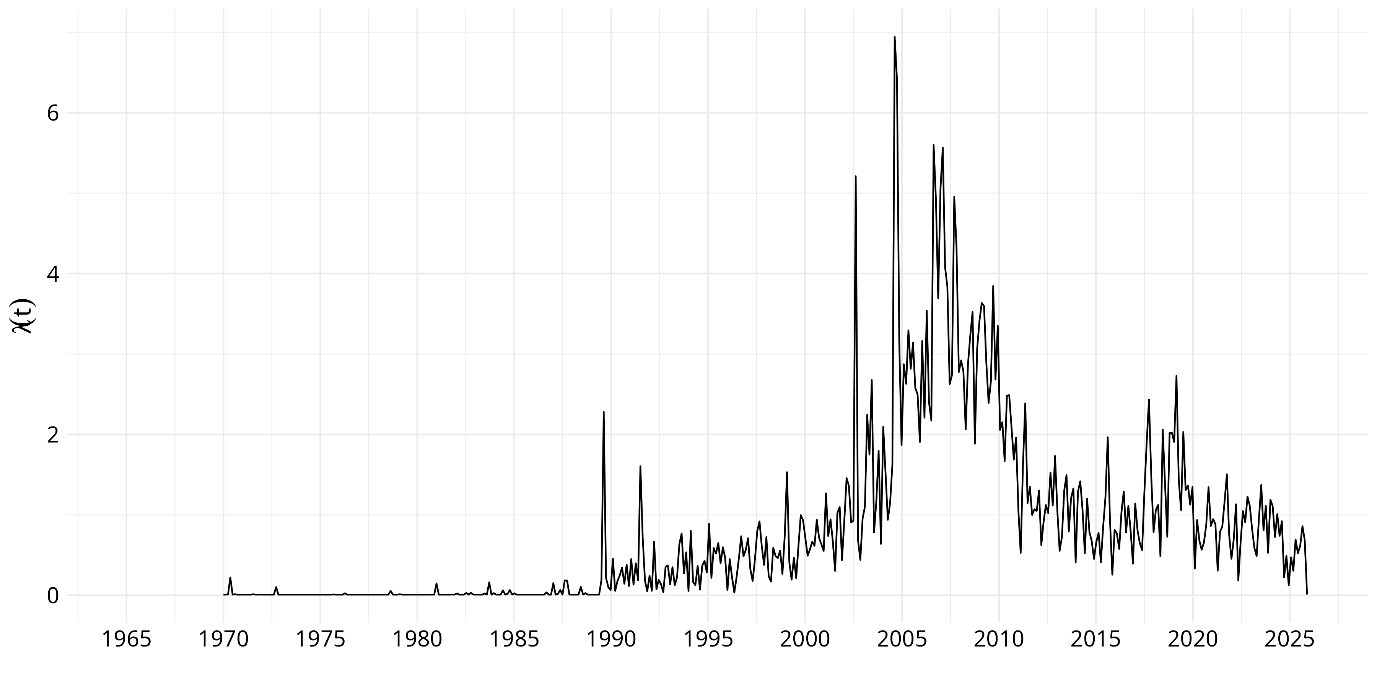

**b)**

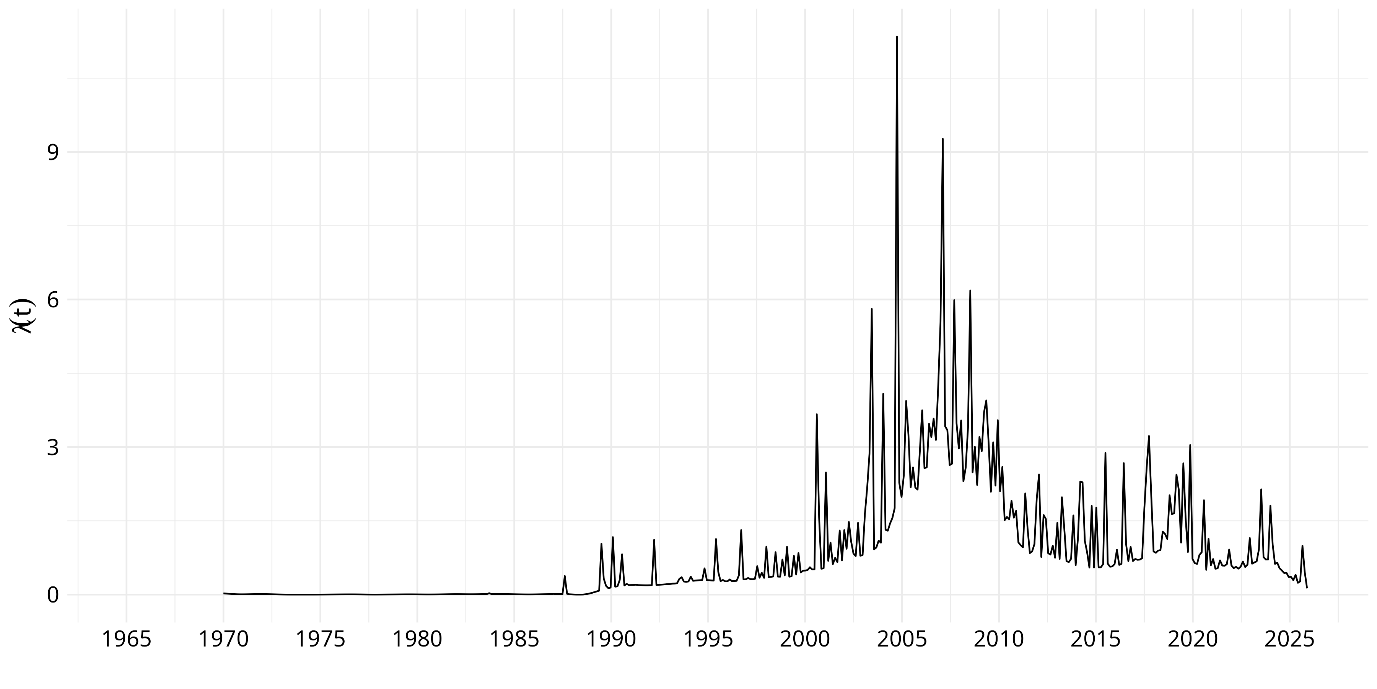

**Figure S7**. Hawkes process models of sexual violence events in the Colombia armed conflict from 1964-2024 with **a)** constant baseline **b)** adjusted for base rate

**a)**

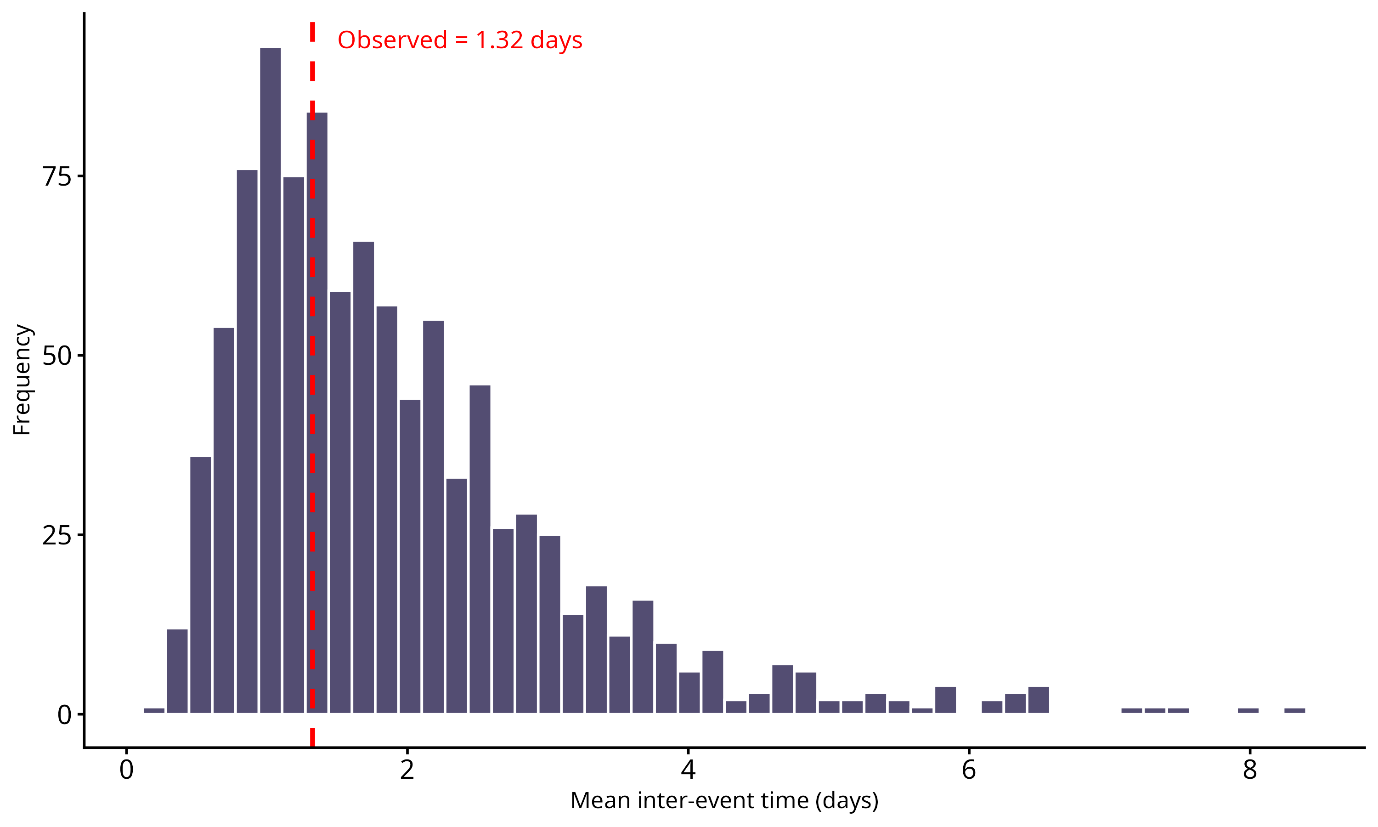

**b)**

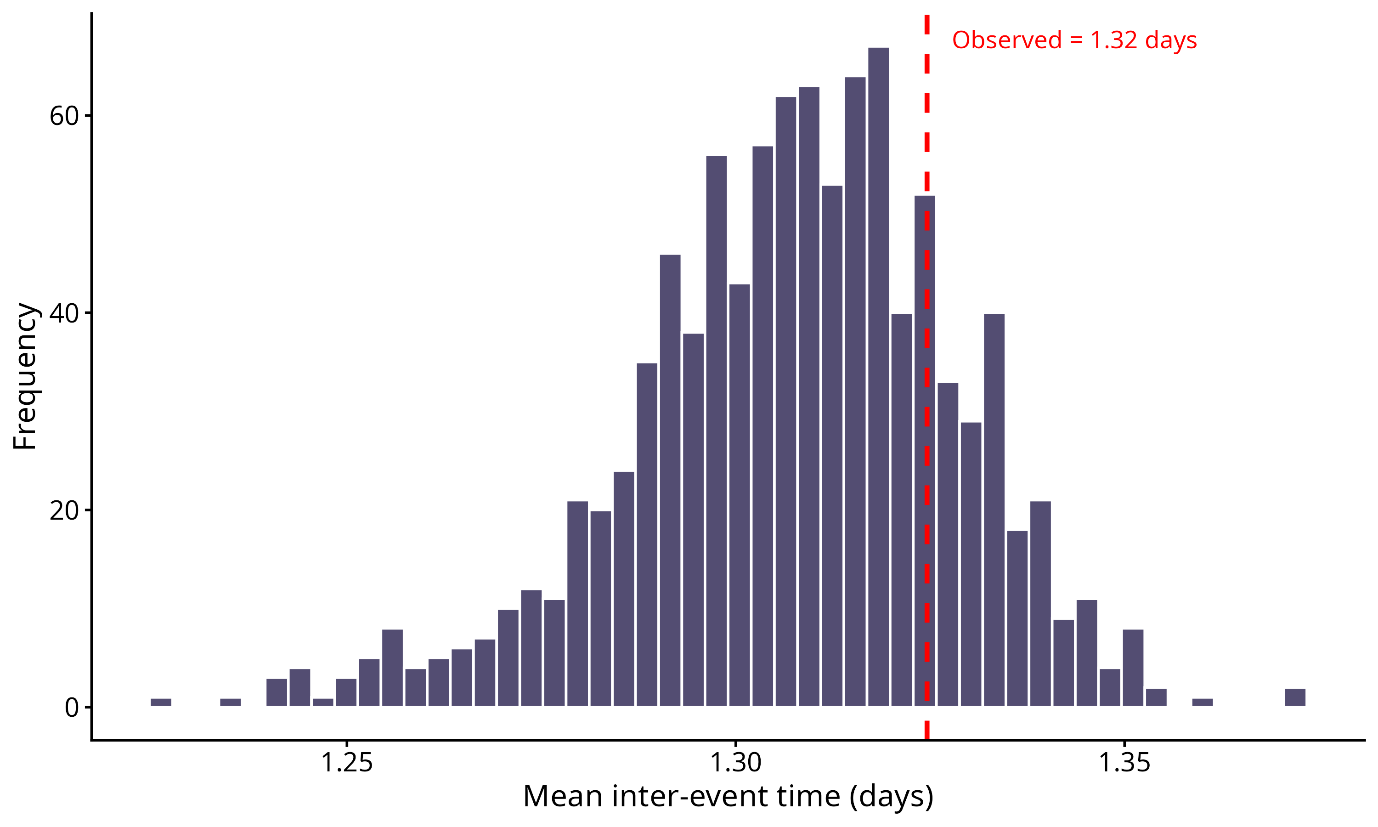

**Figure S8**. Plot of 1000 simulated mean inter-event time from fitted Hawkes process model of sexual violence events in the Colombia armed conflict from 1964-2024 with **a)** constant baseline **b)** adjusted for base rate

**a)**

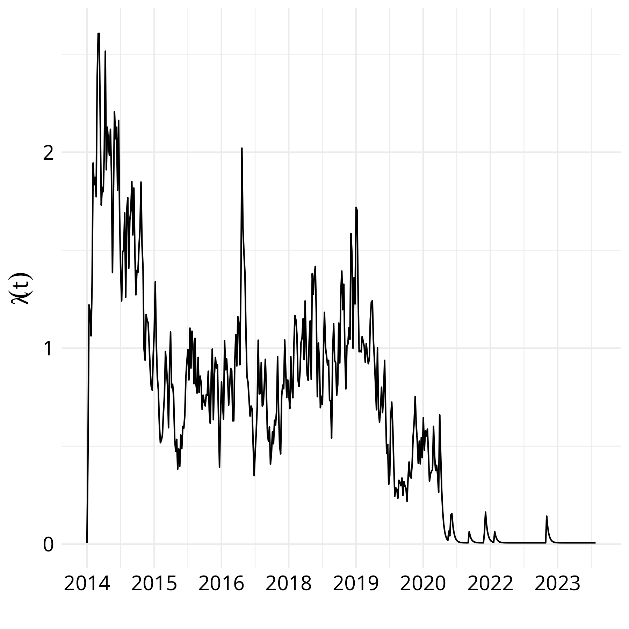

**b)**

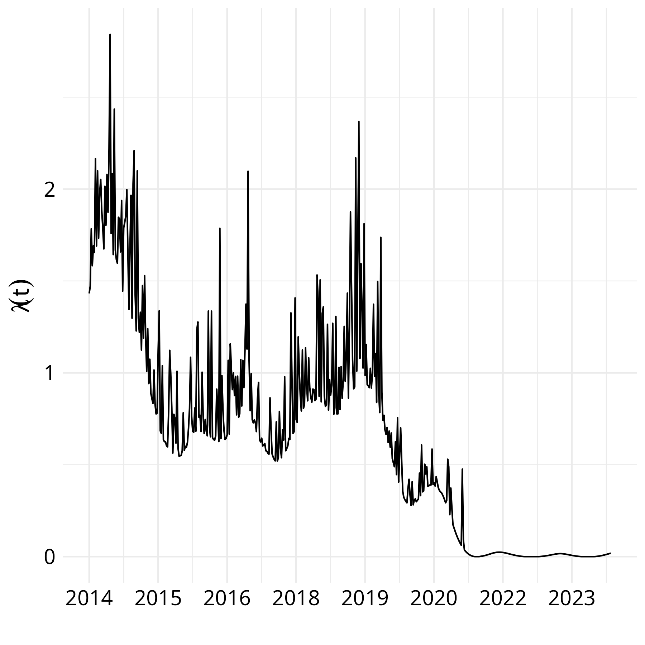

**Figure S9**. Hawkes process models of sexual violence events in the Colombia armed conflict from 2014-2024 with **a)** constant baseline **b)** adjusted for base rate

**a)**

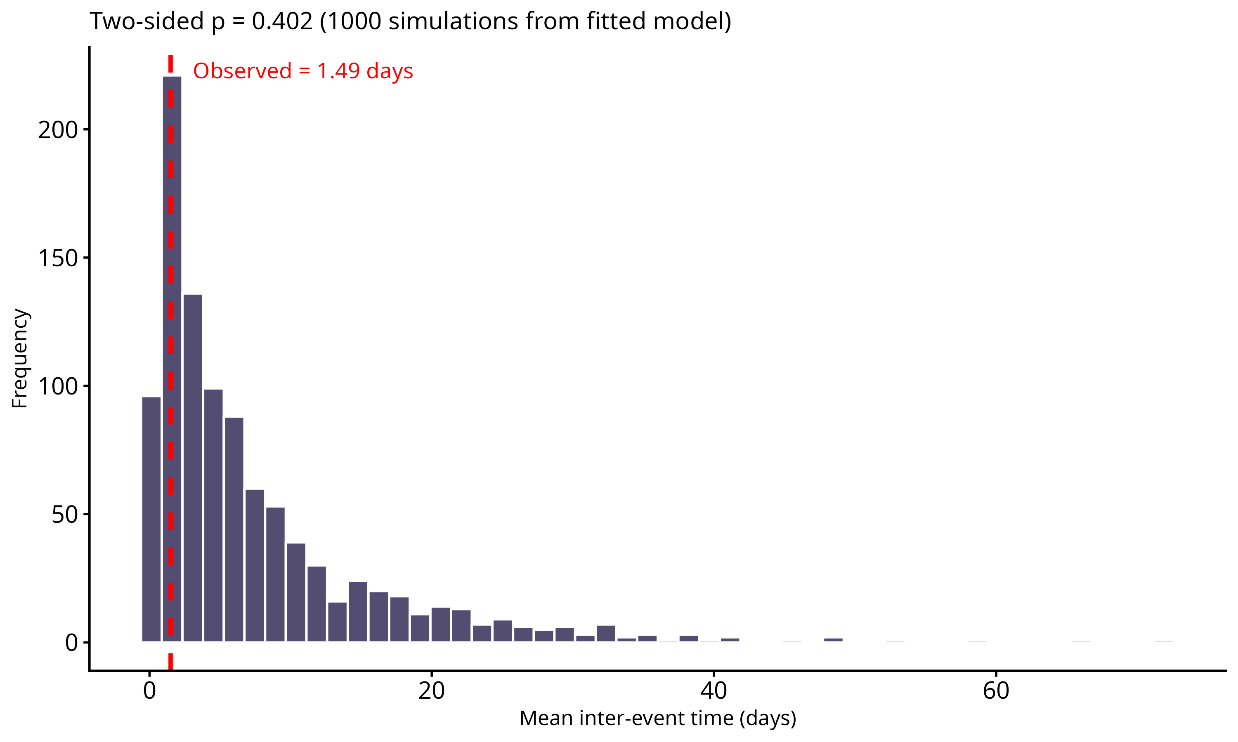

**b)**

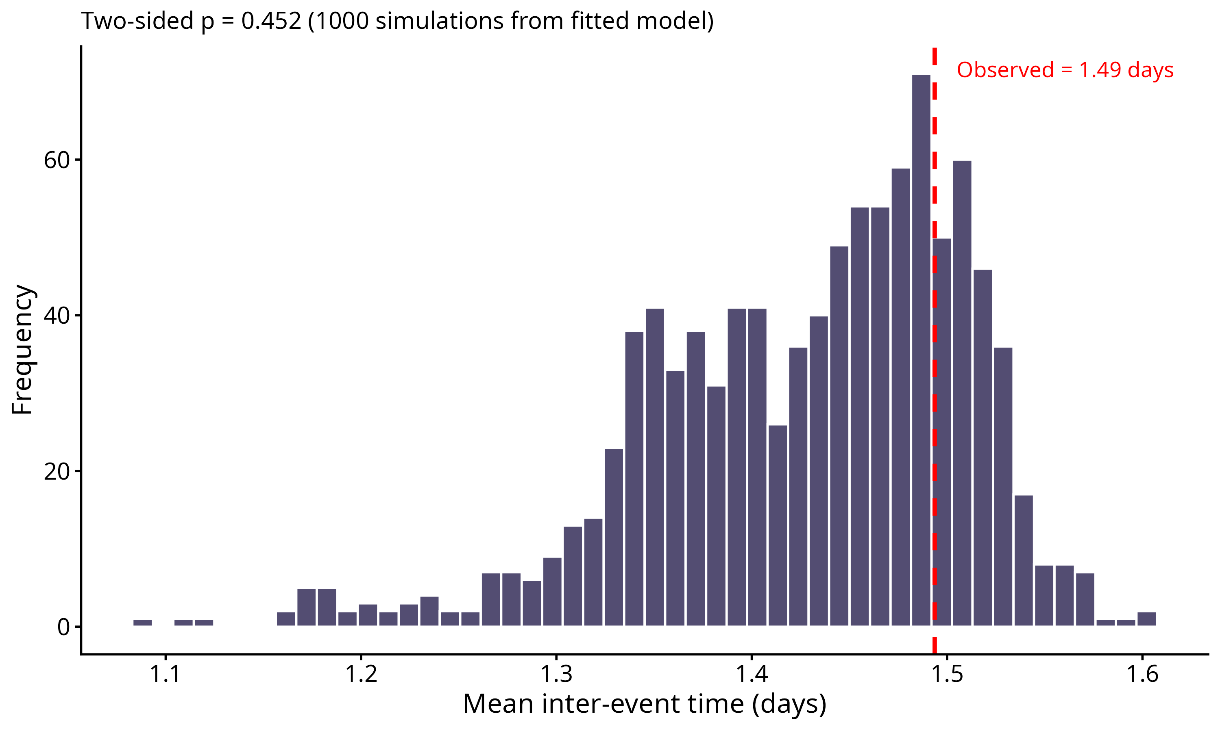

**Figure S10**. Plot of 1000 simulated mean inter-event times from fitted Hawkes process model of sexual violence events in the Colombia armed conflict from 2014-2024 with **a)** constant baseline **b)** adjusted for base rate

**a)**

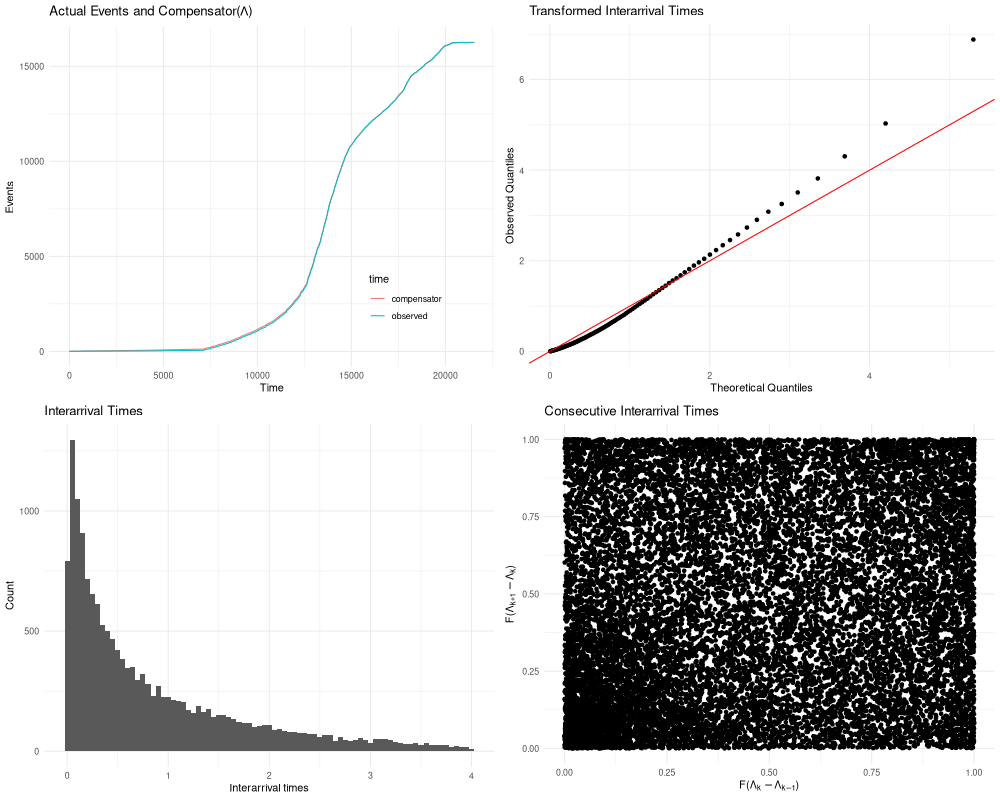

**b)**

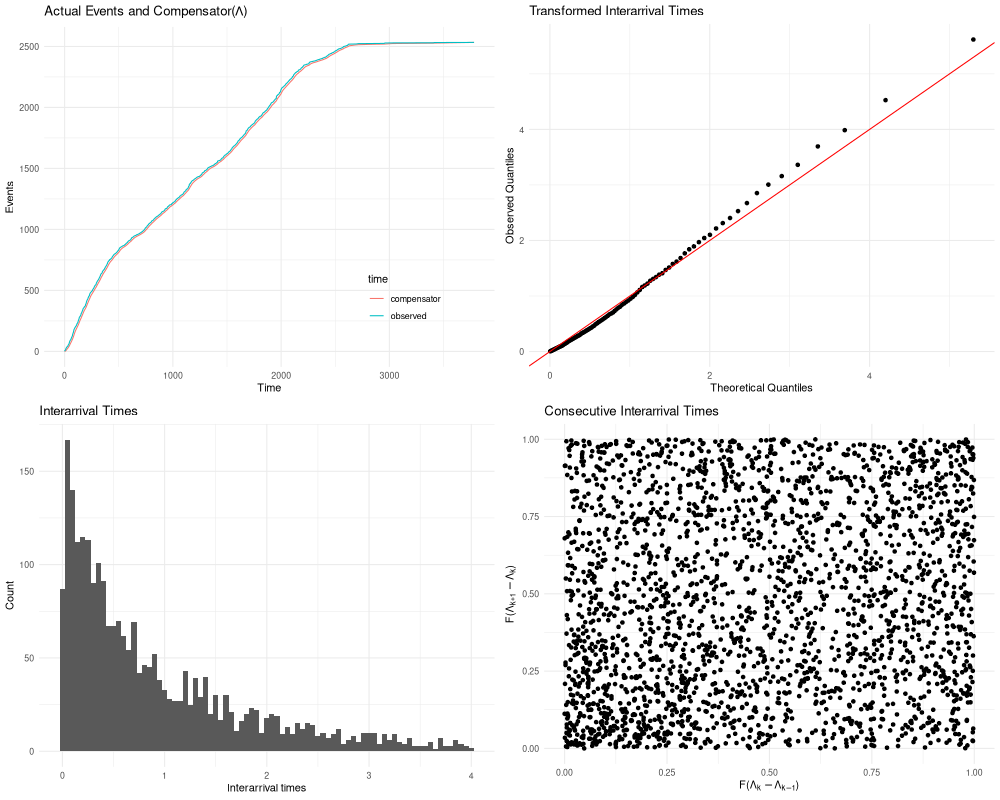

**Figure S11**. Plot of Hawkes process model goodness of fit metrics for Colombia armed conflict data from **a)** 1964-2024 **b)** 2014-2024
